# Beyond influenza-like-illness: data-driven syndromic surveillance for fine-scale monitoring of seasonal respiratory viruses

**DOI:** 10.64898/2025.12.03.25341530

**Authors:** Ronan Corgel, Andrew Tiu, Shweta Bansal

## Abstract

Effective seasonal respiratory virus surveillance in the United States is critical to minimize healthcare burden and maintain economic productivity. Syndromic surveillance aims to monitor disease trends based on symptom occurrence but suffers from outbreak identification issues since case definitions are not disease-specific. We leverage a high-volume medical claims database, representing over 40% of the U.S. population annually from 2016-2023, to develop data-driven syndromic profiles for both influenza and RSV with a regression modeling approach. We then apply these syndromic profiles to estimate total symptomatic disease dynamics and burden by geography and demography. Symptom-inferred disease estimates show strong agreement with existing surveillance systems for both spatiotemporal trends and disease burden. Across all seasons examined, influenza prevalence was largely spatially homogeneous at the county-level, with consistency between seasons and age groups. Data-driven syndromic surveillance can provide fine-scale, representative disease tracking that strengthens forecasting, informs resource allocation, and enhances public health preparedness.

## Introduction

In the United States, seasonal respiratory viruses including influenza and respiratory syncytial virus (RSV) are responsible for millions of infections each year that put significant burden on the health-care system (CDC Influenza Burden Estimate, CDC RSV Burden Estimate, CDC COVID-19 Burden Estimate). Effective surveillance of these pathogens informs critical public health functions such as forecasting hospitalizations and identifying demographic groups to prioritize for vaccination [1, 2, 3]. While strong surveillance systems improve outbreak preparedness, respiratory virus monitoring is not a simple task. For cases to be included in traditional surveillance systems, individuals must seek care at a participating medical facility, receive a positive test result for infection, and have their result reported to a public health agency. Thus, the true dynamics and burden of common respiratory infections are poorly understood, especially at local geographic scales.

Syndromic surveillance examines patients’ symptoms and clinical signs to estimate otherwise unobservable disease dynamics [4, 5, 6]. By focusing on the symptoms associated with infection, syndromic surveillance has potential to estimate true disease due to the infeasibility of conducting population-wide laboratory testing. This type of surveillance for respiratory infections gained traction during the 1990s, however, significant investments in the practice occurred due to bioterrorism threats following the 2001 World Trade Center and anthrax attacks [7, 8, 9]. Identifying opportune data streams, validating symptom sets against respiratory conditions, and evaluating signal timing synchronies were early challenges of syndromic surveillance. Previous research aimed to address these issues by examining a variety of data sources from preclinical to emergent settings, confirming the respiratory syndromes associated with virologic test positivity, and demonstrating that syndromic signals have potential to act as leading indicators of disease [10, 11, 12]. Despite these findings, there remains a key disconnect between syndrome and disease. Many syndromic surveillance systems suffer from identification issues and false positive alerts as syndromic definitions are not specific to disease, instead identifying a broad spectrum of infections [4, 13, 14, 15, 16].

Due to its intense seasonal patterns and pandemic potential, influenza is a primary focus of seasonal respiratory disease surveillance. Influenza symptomatology includes respiratory symptoms (e.g., cough), systemic symptoms (e.g., fever, fatigue, myalgia), and gastrointestinal issues (e.g., nausea and diarrhea) (CDC Influenza Symptoms) [15, 17]. It is common for influenza to be monitored with syndromic surveillance, based on detecting influenza-like illness (ILI), often defined by symptoms such as fever, cough, or sore throat. The European Influenza Surveillance Network (EISN) collates sentinel ILI data from over 30 European countries, while the US Centers for Disease Control and Prevention (CDC) monitors ILI visits through a network of physicians (ILINet), where approximately 4,000 outpatient healthcare providers (out of over 50,000 nationwide) submit reports on the percentage of patient visits with ILI weekly throughout the year (CDC Influenza Surveillance, (Outpatient Healthcare Providers) [18]. Previous work has aimed to validate the surveillance of influenza-like-illness, however, the case definition has consistently been found to correlate with multiple respiratory infections and provide low accuracy in diagnosing influenza itself [11, 16, 19].

More traditional methods of influenza monitoring in the United States involve virologic testing surveillance and laboratory-conformed influenza hospitalization surveillance, a system referred to as FluSurv-NET. Multiple measures from both syndromic and traditional data sources are used to characterize influenza incidence like the percent of outpatient visits categorized as ILI and the influenza hospitalization rate. Each of these measures are available at large geographic scales such as the national or state level, far from the local scale that influenza is known to heterogeneously spread [20, 21, 22, 23]. To estimate influenza prevalence, the CDC utilizes FluSurv-NET data, including roughly 9% of the population across only 14 states (CDC Influenza Hospitalization Surveillance). Not only is this a restricted sample, but the burden estimate is highly dependent on clinicians’ testing decisions. While there have been major improvements in the United States’ influenza surveillance infrastructure in the last few decades, current limitations include narrow representation and a lack of geographic granularity in influenza monitoring.

In the era of big data, opportunistic information sources such as administrative healthcare data can enhance timeliness, spatiotemporal granularity, and representativeness of surveillance compared to traditional disease monitoring systems [24]. One such source that offers high coverage and detailed disease information is medical claims. Medical claims are billing documents produced for every clinical visit in the United States and submitted to insurance companies for reimbursement. Although these data are administrative in nature, patient information, such as disease diagnoses and symptom presence, is recorded. Past work examining medical claims data has demonstrated their usefulness for disease surveillance, characterizing transmission patterns, and obtaining vaccine coverage estimates [21, 22, 25, 26, 27, 28]. In particular, this research found that disease signals from medical claims align with traditional data sources, but showcased the biases created by spatial aggregation and limited reporting. Furthermore, recent research has even examined the ability of medical claims to accurately estimate reported COVID-19 hospitalizations in real-time [29]. While medical claims research has improved our fundamental understanding of disease dynamics and surveillance gaps, past findings rely on tested diseases which are subject to clinician decision behaviors and therefore do not reflect true burden.

Providing accurate respiratory virus surveillance that enables effective policy-making is an ongoing challenge. Where syndromic surveillance is independent of diagnostic testing and more easily scaled to large groups, current syndrome definitions are not disease-specific. Conversely, traditional surveillance methods are superior at detecting individual pathogens, but they cannot test entire populations for infection. Together, each surveillance type struggles with comprehensive geographic coverage and representativeness, which clouds respiratory disease incidence and prevalence estimates for the United States. In this work, we aim to address these surveillance trade-offs by developing data-driven and disease-specific symptom profiles for fine-scale syndromic surveillance of respiratory viruses, with influenza and RSV as case studies. With our tailored disease definitions, we estimate spatiotemporal infection patterns at the county-week scale, providing fine-scale and representative surveillance. We then scale up these estimates to infer unobserved symptomatic influenza prevalence for the general population and specific age groups, finding strong agreement with formal CDC burden estimates. Extending this approach to the county level reveals previously uncharacterized local variation in influenza prevalence. Through our work, we demonstrate the accuracy of local symptom-based disease estimates from administrative healthcare data and provide a rationale for a data-driven syndromic surveillance system in the United States. Following successful implementation, such a surveillance system can monitor previously unobserved disease at fine spatiotemporal scales, improving situational awareness, intervention planning, disease forecasting, and overall public health preparedness for future respiratory disease threats.

## Methods

### Study design

The goal of this study was twofold: First, to create data-driven syndromic definitions of disease and second, to examine the dynamics of those symptom-based definitions at the population-level. We analyzed a large electronic medical claims database covering all 50 US states and the District of Columbia from September 2016 until August 2023. Using these data, we developed statistical models that predicted a patient’s disease status from their symptom profile, demographic information, and population-level disease. From model predictions, we compared spatiotemporal trends and disease burden estimates against confirmed case data and existing disease surveillance sources for validation purposes. We then used the validated models to estimate county-level disease incidence and prevalence. We focus on influenza as a primary and RSV as a secondary case study to demonstrate generalizability.

### Medical claims data

Medical claims are billing documents that traditionally serve as a record for insurance reimbursement purposes [27]. These data were provided by a medical claims clearinghouse that acts as an intermediary between healthcare providers and insurance companies. Across geographies and demographic groups, patients represented in the medical claims accounted for approximately 40% of the United States population (Medical Claims Data Descriptor). Individuals included in the data were those who sought inpatient or outpatient medical care at a provider in the clearinghouse network and possessed private or public insurance at the time of the visit. Data on confirmed influenza and RSV infection diagnosis, as well as symptom presence, were determined using the International Classification of Diseases 10th revision (ICD-10-CM) diagnosis codes (Tables S1, S2, ICD-10). Symptoms recorded were fever, cough, sore throat, bronchitis, fatigue, myalgia, headache, congestion, hypoxemia, chest pain, shortness of breath, diarrhea, nausea/vomiting, and sneezing for influenza; and fever, cough, sore throat, bronchitis, fatigue, headache, congestion, hypoxemia, loss of appetite, shortness of breath, and sneezing for RSV [15, 17, 30]. Medical visit week, U.S. county of residence, age group, and gender information were also extracted from the claims data. Patient counts were inferred by county of residence, week of visit, demographics, confirmed disease status, and symptom profile. U.S. counties with populations smaller than 20,000 were grouped with neighboring counties. To disaggregate the grouped counties, counts were allocated to counties proportionate to their population size. Patient group counts between 1 and 5 individuals were censored, and were imputed by assigning a random number between 1 and 5 to the censored value.

### External surveillance data

Official surveillance data sources were included in the analysis both for comparison and as inputs to predictive models. State-level and national-level influenza virologic data were provided by the National Respiratory and Enteric Virus Surveillance System (NREVSS), representing laboratory tests performed in both inpatient and outpatient settings. Data on laboratory-confirmed influenza hospitalizations were accessed through the Influenza Hospitalization Surveillance Network (FluSurv-NET), which covers 14 U.S. states. For data with increased spatial granularity, New York State (NYS) influenza laboratory-confirmed cases were examined at the U.S. county level to compare with model predictions. These data include test results reported from participating clinical laboratories in New York State. Aside from laboratory-confirmed influenza, data from the Influenza-Like Illness Surveillance Network (ILINet), a sentinel syndromic surveillance system of over 4,000 outpatient clinics, were included to determine model agreement with a syndromic signal defined by the presence of fever and cough and/or sore throat. National influenza prevalence estimates came from the Centers for Disease Control and Prevention (CDC). Finally, wastewater surveillance data from WastewaterSCAN were used by combining time series of influenza A and influenza B concentrations (additional details in Supplementary Information).

### Learning disease-specific symptom profiles

A sample of medical claims data were used to train and test a statistical model, learning disease-specific symptom profiles. The dependent variable was confirmed disease status (either for influenza or RSV) while independent variables included a patient’s symptoms (one of 14 for influenza and one of 11 for RSV), their demographics, and population-level disease estimates. Data spanned September 2016 to August 2020 and the dataset was limited to symptomatic visits for patients with a confirmed diagnosis and those without. A random, balanced sample was taken across time and space, resulting in 500,000 cases and 500,000 non-cases for each disease examined. Since patients had potential to be sampled more than once, we tested the independence assumption of our models by fitting to deduplicated data as well and confirmed little change in results (Figure S1).

With the balanced training dataset, we fit logistic regression models to the influenza and RSV samples. Each model iteration included additional information to examine performance improvements from supplementary data on symptom presence, age group, gender, and population-level disease. For influenza, model 1 (Fever model) only used a binary fever indicator to predict influenza diagnosis of an individual. Model 2 (Symptom model) incorporated all symptom variables as binary indicators in our dataset. Model 3 (Interaction model) included demographic information along with all symptoms to predict disease status. This model also included interaction terms between symptoms and age group as well as symptoms and gender to account for known differences in immunology and symptom presentation by age and sex [31] [32]. Model 4 (Case model) added the weekly county-level proportion of visits with confirmed influenza lagged by one week to model 3. In model 5 (NREVSS nodel), we alternatively added the weekly influenza state-level virologic data from NREVSS lagged by one week to model 3. In instances where state-level test positivity was missing, national-level data was used. model 6 (Wastewater model) added wastewater data to model 3, but only for 2022-2023, when wastewater data were available. While previous models were fit using a logistic generalized linear model (GLM), the Case, NREVSS, and Wastewater models utilized logistic generalized additive models (GAMs) to allow for flexibility in the relationship between the population-level disease estimate and visit-level disease status. All models were fit for both influenza and RSV, with the exception of the NREVSS and Wastewater models which were only fit for influenza due to data availability. We evaluated model performance using 10-fold cross-validation. The receiver operating characteristic (ROC) and corresponding area under the curve (AUC) were estimated for each model and compared to determine predictive ability. Final versions of each model were trained on the complete, balanced sample of data following cross-validation. We chose classification cutoff values by maximizing the Youden index, calculated as sensitivity + specificity - 1. Optimal cutoff values varied by age group, so age-specific cutoffs were utilized for the remainder of the analysis.

### Predicting care-seeking symptomatic disease incidence from syndromic models

Models trained and tested on sampled, balanced data were applied to the full, unbalanced, medical claims dataset, creating disease-specific estimates based on syndromic dynamics. Aggregated patient information on symptom occurrence at the county-week scale provided the basis for predictions of symptomatic disease, assuming a geographically and temporally homogeneous relationship between symptoms and disease. We tested this assumption and found general consistency in the disease-symptom relationship between states and across months (Figure S2). Data were aggregated to the county, state, and national levels to investigate disease timeseries and estimate key seasonality metrics. Timeseries data were smoothed at each respective spatial scale with a moving average encompassing the current week, two preceding weeks, and two future weeks. We defined season peak time as the largest local maximum exceeding both 10 cases and 2.5 standard deviations above the summer mean incidence (a threshold determined empirically from confirmed influenza timeseries), selecting the earlier date in the case of ties.

### Estimating unobserved symptomatic influenza prevalence across seasons and age groups

We estimated population-level symptomatic influenza prevalence, extending beyond care-seeking cases to infer disease burden regardless of healthcare-seeking or testing. Predicted care-seeking symptomatic weekly incidence was summed by county from October 1 through April 30 (the official respiratory virus season) to obtain total care-seeking symptomatic influenza visits per season. We then scaled these counts up to account for healthcare-seeking behavior, using the established range that 42% to 58% of symptomatic influenza cases seek care [33], applied as a uniform distribution. Scaled case counts were normalized by total healthcare visits during the same period to estimate prevalence by season. Finally, we corrected for geographic differences in testing rates and medical coding practices by multiplying county-level burden estimates by the ratio of the national to county-level influenza testing rate, downweighting counties with high testing rates and upweighting those with low testing rates. We validated our national prevalence estimates against CDC estimates of unobserved symptomatic influenza burden (no similar estimates are available for RSV). We then leveraged our validated approach to produce county- and age-specific prevalence estimates not available from existing surveillance sources.

### Code availability

All analyses were conducted in R and code is available through GitHub.

## Results

### Symptoms and confirmed cases follow similar spatiotemporal trends

National trends for influenza and RSV healthcare visits followed clear seasonal patterns, typically peaking during the winter months. Notably, influenza peaks tended to occur in late January and February while RSV peaked earlier in December and the beginning of January (Figures 1a, 1b). Healthcare visits where fever and cough were indicated as a symptom also exhibited seasonality, peaking at a similar time as influenza (Figures 1c, 1d). Other symptoms such as fatigue and myalgia did not show signs of seasonality (Figures S3h, S3m). While influenza- and RSV-associated health-care visits decreased significantly during the summer months, symptom visits leveled off at a higher summer baseline, likely due to the presence of other pathogens and non-infection-related sources. Across diseases and symptoms, the highest prevalence of the condition was often concentrated in the Southern United States during the 2018-2019 respiratory virus season (Figure S4). Throughout the country, the prevalence of symptoms such as fever, cough, and sore throat were much higher than the prevalence of confirmed disease.

**Figure 1:**
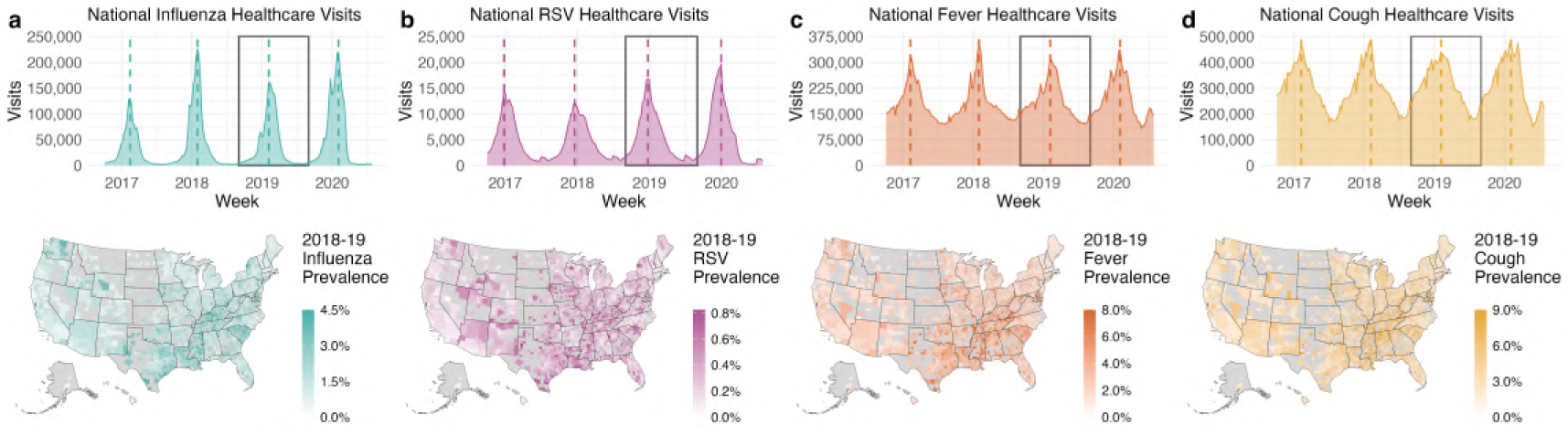
Disease and symptom observed spatiotemporal trends, 2016-2020. (**a**) Confirmed influenza healthcare visits in the United States and county-level confirmed influenza prevalence for the 2018-19 respiratory virus season. (**b**) Confirmed RSV healthcare visits in the United States and county-level confirmed RSV prevalence for the 2018-19 respiratory virus season. (**c**) United States healthcare visits where fever was indicated as a symptom and county-level fever prevalence for the 2018-19 respiratory virus season. (**d**) United States healthcare visits where cough was indicated as a symptom and county-level cough prevalence for the 2018-19 respiratory virus season. Prevalence estimates were calculated by dividing the deduplicated number of disease or symptom cases by the total number of unique patients (all causes) in each county from October 2018 through April 2019. Counties with small population sizes (< 10,000), low numbers of claims (< 5,000), or with outlier values (> 3 standard deviations above the mean) were excluded from the prevalence estimates.

### Symptoms and demographics are predictive of disease

We developed data-driven syndromic definitions for influenza and RSV using medical claims. With medical claims from 2016 to 2020, the proportion of patients coded with influenza-like symptoms varied widely between confirmed influenza cases and non-influenza cases. For instance, 46.0% of influenza cases were coded as having a fever compared to 9.2% of non-influenza cases (Figure 2a). Certain generalized symptoms, such as chest pain and fatigue, occurred more frequently in non-influenza cases than in influenza cases (13.3% vs. 4.6% and 16.0% vs. 7.2%, respectively). RSV exhibited similar patterns, and there were distinct differences between RSV and influenza syndromic profiles (Figure 2c). For example, although sore throat was more common in confirmed influenza cases than in non-cases (21.1% vs. 10.8%), the symptom was more prevalent in non-RSV cases than RSV cases (15.2% vs. 5.1%).

**Figure 2:**
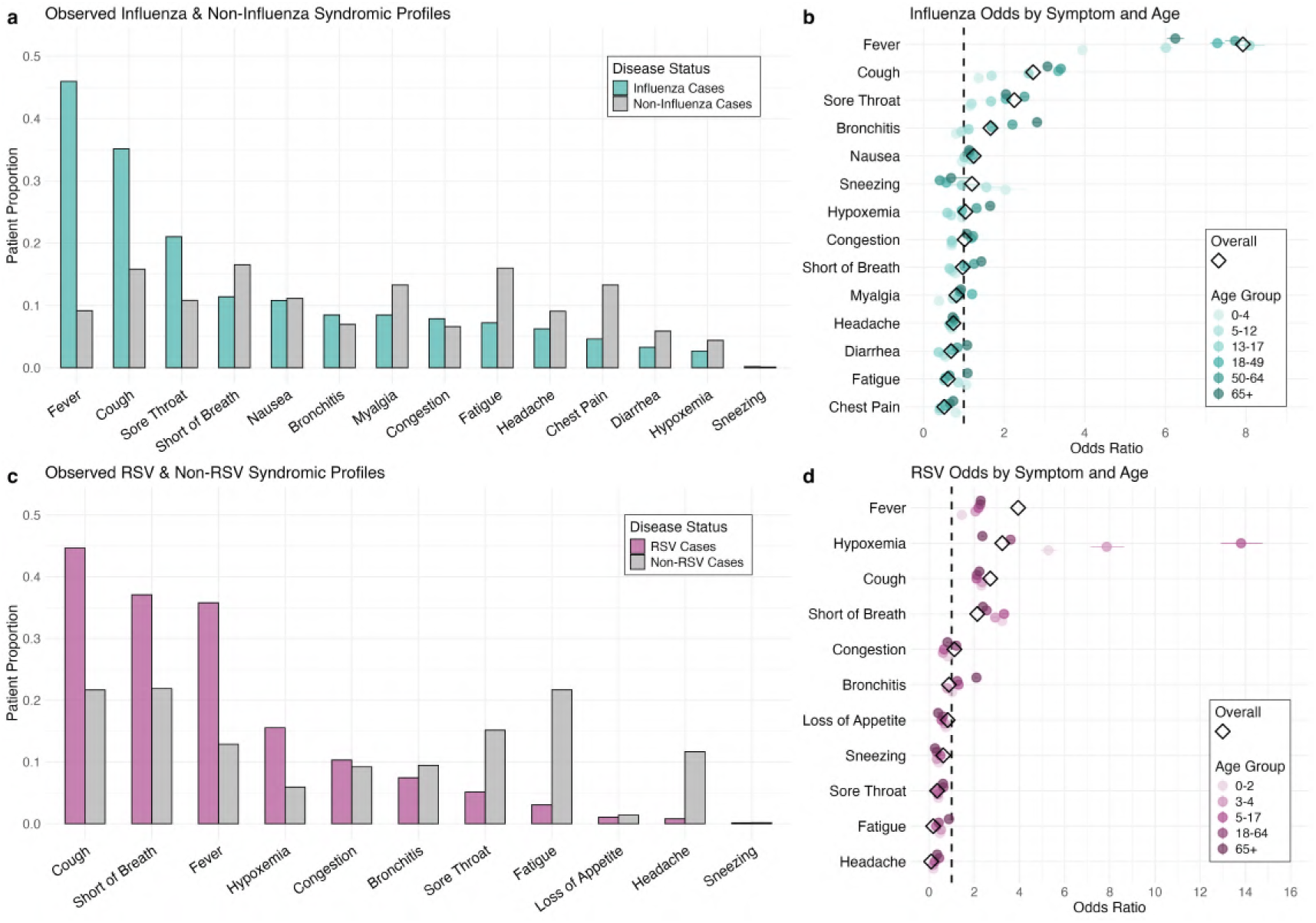
Visit-level symptom occurrence and predictability for influenza and RSV. (**a**) Proportion of influenza cases and non-influenza cases with 14 different symptoms. (**b**) Logistic regression results displaying the odds ratios of influenza given symptom presence, for all ages and by age group. (**c**) Proportion of RSV cases and non-RSV cases with 11 different symptoms. (**d**) Logistic regression results displaying the odds ratios of RSV given symptom presence, for all ages and by age group. Both observed proportions and regression results were produced from balanced samples of cases and non-cases from 2016-2020, representing a subset of the full medical claims data source. Overall and age-specific odds ratios were from the Symptom Model, where confirmed disease was the dependent variable and all symptoms served as independent predictors.

Overall, logistic regression results confirmed symptom predictability observed in the data. In our model containing all identified symptoms for each disease (Symptom model), the odds of influenza were 7.9 times as high in fever patients compared to patients with no fever, and the odds of influenza in chest pain patients were 0.5 times the odds in patients experiencing no chest pain (Figure 2b). Likewise, the odds of RSV were more than twice as high in patients with shortness of breath compared to patients with no breathing difficulty, and the odds of RSV in sore throat patients were 0.4 times the odds in patients not exhibiting a sore throat (Figure 2d). Notably, these odds ratios varied across age groups. While the presence of fever increased the odds of influenza across all age groups, young and old age groups experienced lower odds than middle-aged adults. Similarly, bronchitis increased the odds of influenza in older age groups but not in patients aged 0 to 12 years old. Gender did not substantially modify the relationship between symptoms and disease (Figure S5). Across candidate models, cross-validated AUC values ranged between 0.68 and 0.88 for influenza and from 0.62 to 0.93 for RSV (Figures S6a, S7a). Additionally, we trained and evaluated the same models on data from 2021 to 2023, after the emergence of SARS-CoV-2. We found that across all models for both diseases, AUC values slightly decreased, ranging from 0.65 to 0.80 for influenza and from 0.60 to 0.92 for RSV (Figures S6b, S7b). Regardless of data time period, the Case model proved to be the best-fit for predicting disease for both infections, with the NREVSS model as a close second.

### Syndromic surveillance estimates spatially heterogeneous disease peak times

We applied our fitted symptom models to individual-level claims data across all counties, age groups, and weeks, aggregating predictions to characterize population-scale disease dynamics. We compared predicted population-scale symptomatic disease dynamics against population-scale confirmed case dynamics from medical claims to estimate model performance. The county-level peak time for confirmed cases and predicted syndromic cases was estimated and contrasted across five respiratory virus seasons to determine goodness-of-fit. We found strong agreement in peak time between the best-fit model (Case) and confirmed influenza, with a Spearman correlation of 0.82 and a root mean square error (RMSE) of 12.2 days during the 2016-17, 2017-18, 2018-19, 2019-20, and 2022-23 respiratory virus seasons (Figure S8). The influenza NREVSS model also had high concordance with a Spearman correlation of 0.71 and a RMSE of 17.4 days, and the Wastewater model had a Spearman correlation of 0.63 and an RMSE value of 10.0 days (Figure S9c). County-level peak times estimated from the best-fit RSV model produced a 0.28 Spearman correlation and a RMSE of 25.8 days when compared to peak times from confirmed RSV during the same respiratory virus seasons (Figure S10). Influenza symptomatology was more identifiable than RSV, where even the Fever model for influenza outperformed the most complex RSV model.

In general, syndromic predictions from the best-fit influenza model (Case) agreed strongly with traditional influenza surveillance data sources. At the state level, Pearson correlation coefficients between predicted and traditional surveillance time series ranged from 0.26 to 0.96 for ILI-NET, FluSurv-NET, and NREVSS sources (Figure 3a). When comparing state-level peak timing, Spearman correlation coefficients were 0.82, 0.87, and 0.84 for ILI-NET, FluSurv-NET, and NREVSS across respiratory virus seasons, respectively (Figures S11a, S11b, S11c). In addition to state-level agreement, syndromic predictions aligned to external sources at the county-level. Across counties in New York, Pearson correlation coefficients between syndromic predictions and New York State lab-confirmed time series data ranged from 0.11 to 0.88 (Figure 3a). The Spearman correlation coefficient between predicted and New York State county peak times was 0.83, comparable to state-level peak time correlations (Figure S11d). Figure 3b captures a key difference between surveillance sources for the estimation of season duration. ILINet’s broad case definition likely captures non-influenza infections, producing a longer apparent season, while FluSurv-NET’s reliance on laboratory-confirmed hospitalizations yields a shorter, more restricted one. Our syndromic predictions fall between these extremes, balancing specificity and sensitivity by using a disease-specific symptom profile without requiring diagnostic confirmation.

**Figure 3:**
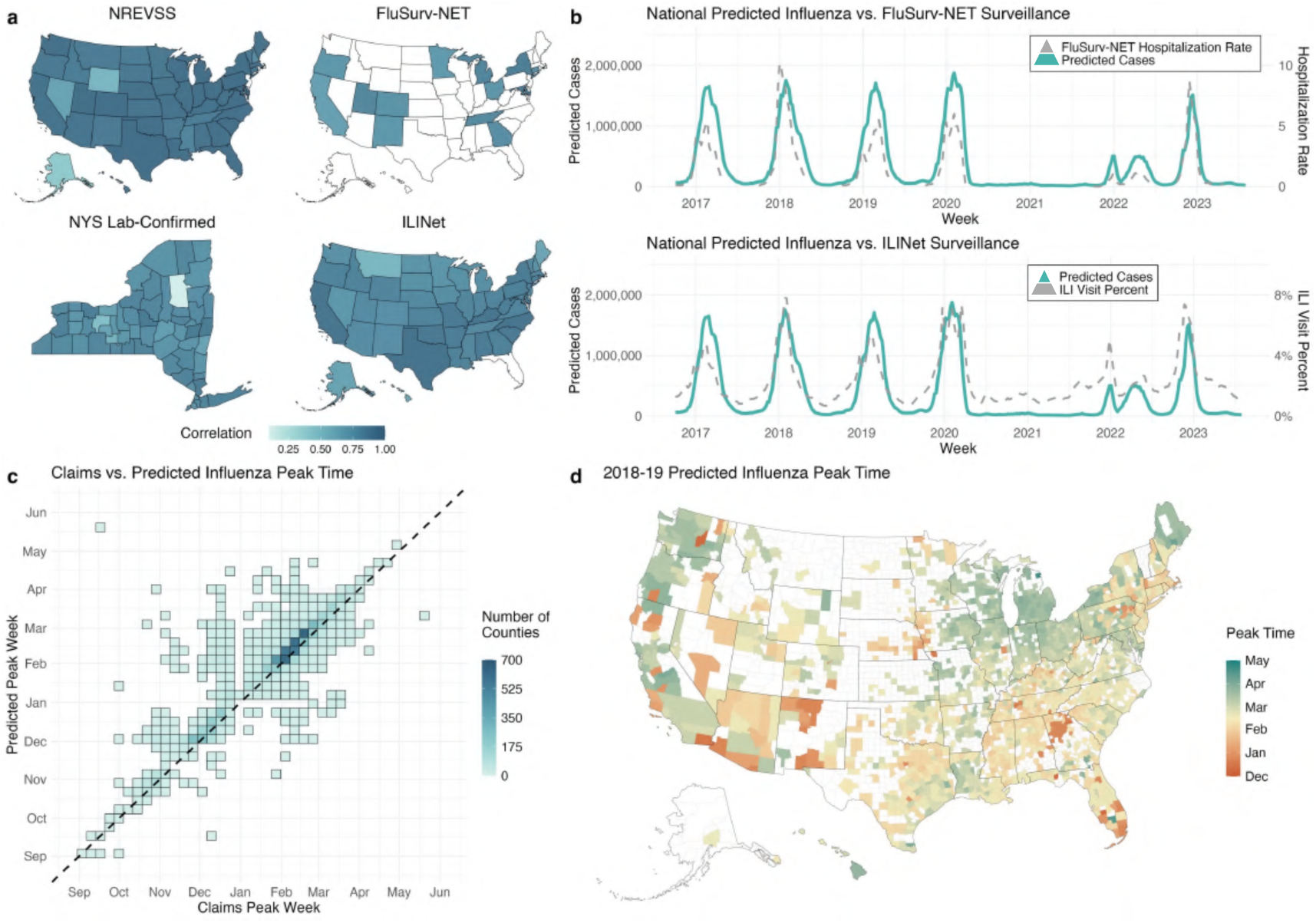
Spatiotemporal validation and county-level prediction of influenza peak times. (**a**) State and county-level Pearson correlation coefficients between predicted influenza cases and external surveillance timeseries. White spatial units represent areas where no data was available for the external data source. (**b**) National predicted influenza case timeseries compared against two external surveillance sources: FluSurv-NET and ILINet. A secondary axis is used to plot the external surveillance source. Pyramid legends describe the specificity of the surveillance source. Since FluSurv-NET tracks lab-confirmed hospitalizations, the system captures fewer cases than syndromic influenza predictions. Conversely, ILINet’s broad case definition likely includes more patients with other conditions than our syndromic influenza predictions. (**c**) County peak time correlation between predicted influenza from the Case Model and confirmed influenza during the 2016-17, 2017-18, 2018-19, 2019-20, and 2022-23 seasons. The 2020-21 season was excluded due to low influenza circulation during the COVID-19 pandemic while the 2021-22 season was excluded due to the occurrence of unusual multi-peak dynamics. (**d**) U.S. county predicted influenza peak times from the Case Model for the 2018-19 respiratory virus season. White counties either represent areas with fewer than 10,000 inhabitants, fever than 5,000 total claims during the season, or where no peak was identified. A five week, centered, rolling average was completed to remove any noise in the data, especially in sparsely populated counties. For each timeseries at all spatial scales, Z scores were calculated by subtracting summer mean weekly incidence between June and August from the weekly incidence value and then dividing by the standard deviation of the entire timeseries for the respiratory virus season.

After validating our results, we described the spatial and seasonal heterogeneity of county-level peak timing for influenza and RSV activity based on our syndromic predictions from the best-fit model of each disease. Influenza county-level peak timings showed strong heterogeneity across both counties and seasons (Figures 3d, S12a). For instance, during the 2018-19 respiratory virus season both the metropolitan areas of Atlanta and Miami experienced early influenza peaks in December relative to the rest of the country. However, for the previous season, these regions saw influenza activity peak in early February, on par with a majority of the United States. The spatial pattern of RSV peak times were more consistent across seasons, where the Southeastern part of the country tended to experience earlier peaks than the Northeast and Western United States (Figure S12b). Strikingly, the 2022-23 respiratory virus season saw historically early peak times for both influenza and RSV throughout the entire country. These findings highlight the importance of spatially explicit surveillance data, since counties within the same state experienced widely different peak times.

### Predicted syndromic influenza prevalence describes unobserved burden at fine scales

Next, we examined our syndromic prediction’s ability to estimate unobserved symptomatic disease prevalence. After accounting for healthcare-seeking behavior, normalizing based on the total seasonal patient visits, and correcting for testing biases, we found that predicted prevalence aligned closely with the formal estimates provided by the CDC. Across seasons and age groups, 95% confidence intervals from our predicted prevalence values consistently overlapped with 95% confidence intervals from the CDC (Figure 4a). For example, our prevalence estimates described higher burden seasons in 2017-18 and 2019-20 with a considerably low burden season occurring in 2021-22, similar to external evidence. During the 2016-17 influenza season, our age-specific estimates were more precise than the CDC estimates, particularly for infants and toddlers, for whom no reliable prevalence estimate exists. Notably, we also predict a higher mean burden in school-age children and adults over 65 than the CDC estimate (Figure 4b). All model versions produced burden estimates similar to those of the CDC, except for the Fever model, which routinely underestimated prevalence (Figure S13).

**Figure 4:**
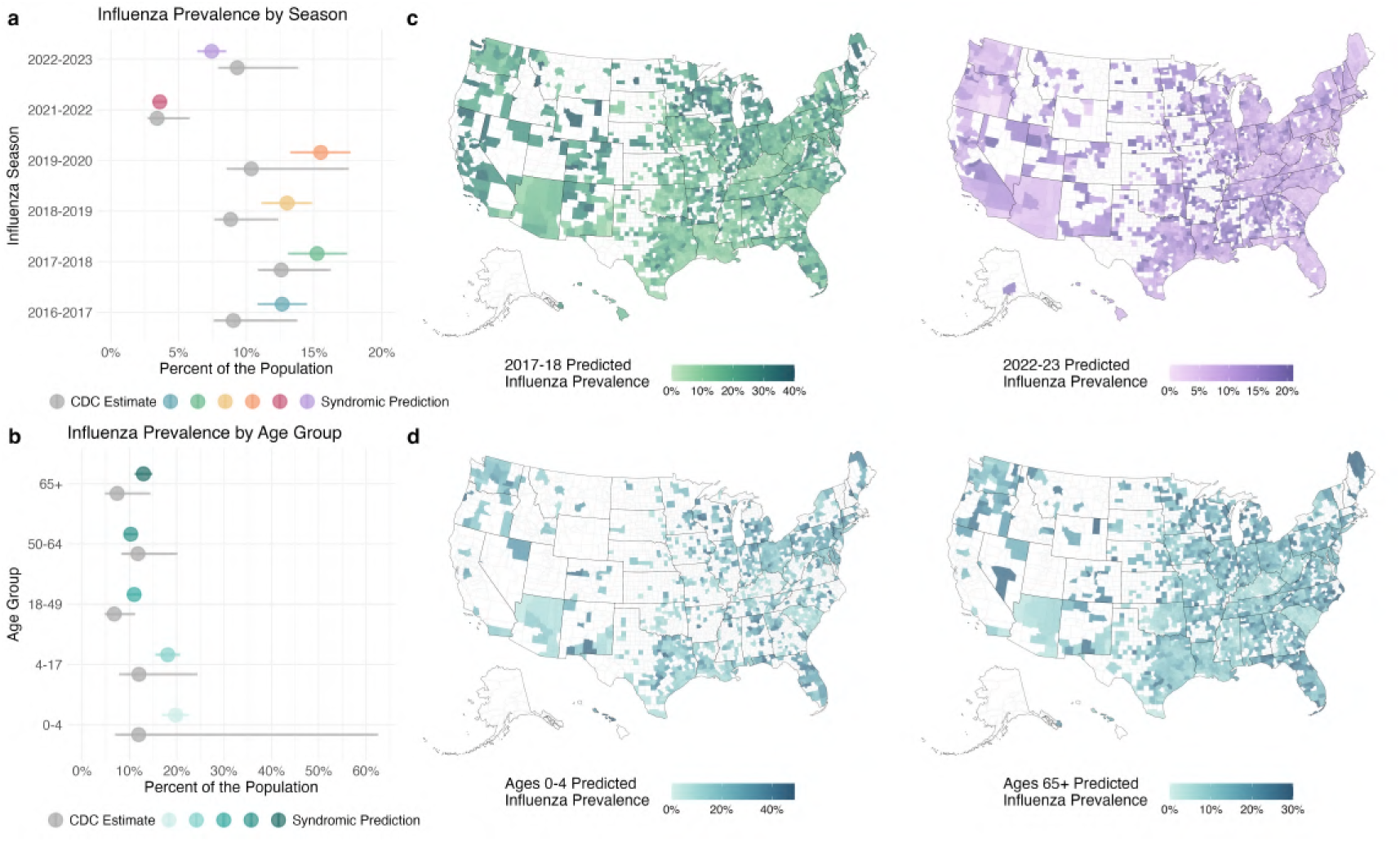
Influenza prevalence validation and county-level estimation by season and age group. (**a**) National influenza prevalence for the entire population comparing the CDC estimate to the NREVSS Model syndromic prediction across seasons. (**b**) Predicted influenza prevalence from the NREVSS Model across counties for the 2017-18 and 2022-23 seasons. White counties either represent areas with fewer than 10,000 inhabitants, fever than 5,000 total claims during the season, or where prevalence estimates were more than two standard deviations above the mean. (**c**) National influenza prevalence for the 2016-17 season comparing the CDC estimate to the NREVSS Model syndromic prediction across age groups. (**d**) Predicted influenza prevalence from the NREVSS Model across counties for the 0 to 4 and 65+ age groups during the 2016-17 season. White counties either represent areas with fewer than 10,000 inhabitants, fever than 1,000 total claims during the season for the age group, or where prevalence estimates were more than two standard deviations above the mean.

The granularity of our data source also allows us to make unobserved syndromic prevalence estimates at the county level. Across seasons, over 50% of county-level influenza prevalence values were between 8% and 20%, demonstrating a largely spatially homogeneous landscape (Figure 4c). Counties with a higher prevalence of influenza during a given season also tended to have high prevalence in other seasons for seasons preceding the emergence of SARS-CoV-2. Post-pandemic seasons (including 2022-23, (Figure 4c)) demonstrate a different spatial distribution of prevalence, as influenza dynamics are still returning to equilibrium. The Spearman correlation coefficient between pre-pandemic seasons ranged from 0.60 to 0.81, while the value was between 0.21 and 0.41 when pre-pandemic and post-pandemic seasons were compared. Within the same season (2016-17) but across age groups, county-level estimates of influenza prevalence were wider for young children than for the elderly (Figure 4d). Spatial correlation between age groups was more pronounced than correlation between seasons, with Spearman coefficient estimates from 0.85 to 0.97.

## Discussion

In this work, we developed data-driven syndromic case definitions for influenza and RSV to create disease-specific symptom profiles of infection. We subsequently validated our models against confirmed cases from medical claims and traditional public health surveillance sources. Since our syndromic models estimated both tested and untested disease, we were able to examine previously unobserved disease dynamics and calculate influenza burden without diagnostic biases. We found that the addition of more symptoms, demographic information, and population-level disease estimates improved model predictability and that influenza was more distinguishable by symptoms than RSV. County-level peak time of predicted syndromic disease activity was spatially heterogeneous and varied across respiratory virus seasons, however RSV exhibited more spatial consistency than influenza. In contrast, we observed overall spatial homogeneity in the predicted influenza burden across U.S. counties. Data-driven syndromic surveillance offers an extensive balance of information, sharing the strengths of traditional public health surveillance and minimizing many of the weaknesses. Our syndromic estimates improve upon the specificity issues of typical ILI surveillance while failing to fall into testing bias and availability pitfalls of traditional surveillance methods.

Syndromic influenza predictions aligned closely to both confirmed cases and traditional surveillance data sources, highlighting the generalizability of the surveillance method. In particular, the Case and NREVSS influenza models performed well during sampled seasons in addition to the out-of-sample 2022-23 respiratory virus season following the introduction of SARS-CoV-2. Despite the emergence of a competing pathogen with overlapping symptoms, peak time and burden estimates continued to agree with confirmed influenza and external sources. RSV was more difficult to capture with syndromic surveillance. Despite high AUC values indicative of adequate model performance, peak time alignment with confirmed RSV cases was worse than influenza. This was likely an artifact of symptom dynamics being representative of influenza rather than RSV, as observed in Figure 1. Future work with multinomial modeling could account for co-circulating pathogens to better distinguish RSV and influenza. Our analysis identified roughly 10 times more symptomatic influenza cases than those with a confirmed diagnosis, consistent with prior evidence of substantial influenza under-detection [34, 33], and in line with estimates from SARS-CoV-2 during the COVID-19 pandemic [35]. While the CDC produces national influenza prevalence estimates each season [36], our approach yields finer spatiotemporal granularity under fewer assumptions and can be applied in near real-time, extending to diseases such as RSV for which formal burden estimates are not routinely produced. Overall, data-driven syndromic surveillance offers a unique opportunity to observe a comprehensive disease landscape in an otherwise fragmented federal public health system.

Our best-performing syndromic models incorporated population-level disease estimates from either medical claims or external surveillance sources, and this information was critical for capturing the seasonal nature of respiratory infections. Without it, predicted summer case counts were inflated, and seasonal patterns were weaker, suggesting models were picking up a broader spectrum of infections rather than a disease-specific signal (Figure S14). This points to an important data fusion principle: individual surveillance sources can combine to produce a system greater than the sum of its parts. That said, the limitations of any external data source are inherited by the model – the wastewater model was restricted to a small number of counties with consistent sampling, and the NREVSS-informed model was constrained by coarse state-level resolution disease estimates. Importantly, symptom data alone can still be informative even without these supplementary inputs: our Fever model showed strong concordance with confirmed influenza peak times, and given that fever has been linked to higher influenza infectiousness [37], fever-based prediction alone can provide a valuable signal for tracking transmission risk.

This study does have limitations arising from simplifying assumptions required for scalability and generalizability. First, medical claims coding practices introduce key limitations. Confirmed diagnosis codes suggest testing occurred, but we cannot verify actual test administration or accuracy. Additionally, symptom diagnosis codes are non-reimbursable, providing little incentive for comprehensive documentation. Recorded symptoms likely represent only a subset of complaints rather than complete clinical presentations, though chief complaints are likely included. For example, previous research found that 13% of secondary influenza cases reported one symptom and 11% were asymptomatic, while in our data 35% of influenza cases had one symptom recorded and 21% were asymptomatic [38]. However, this undercoding is unlikely to systematically bias model estimates, since logistic regression is robust to incomplete covariate data as long as the missingness is not systematically related to the outcome. Second, despite covering insured individuals who represent approximately 92% of the U.S. population, a broader sample than many existing surveillance systems, our medical claims data likely underrepresents vulnerable uninsured populations such as unhoused and undocumented individuals (US Health Insurance Coverage) [39]. Finally, like all influenza surveillance systems, our approach captures only symptomatic cases and thus underestimates total disease burden, which includes asymptomatic cases [40].

Our findings have implications for both clinical practice and public health surveillance. Clinically, disease-specific syndromic profiles enable more accurate case identification than broad ILI definitions, reducing misdiagnosis and improving diagnostic precision at the point of care. At the population scale, these targeted profiles can identify hidden, untested outbreaks while reducing false alarms that are more likely under a typical ILI case definition. The spatial heterogeneity we observed in influenza and RSV peak timing further suggests that geographically uniform guidance applied identically across the entire United States may be insufficient. Vaccination campaigns, antiviral distribution, and risk communication would benefit from locally tailored, spatially explicit disease estimates rather than national or regional averages. This is particularly important given that different respiratory pathogens affect distinct patient populations, require pathogen-specific treatments, and have their own vaccines. Timely, fine-scale burden estimates from data-driven syndromic surveillance could therefore directly inform resource allocation decisions, ensuring appropriate pediatric and adult ICU capacity, sufficient antiviral supply, and well-timed vaccination efforts for the correct pathogen in the right place. Importantly, data-driven syndromic surveillance is not intended to replace traditional disease monitoring but to complement it, filling spatial, temporal, and demographic gaps that existing systems cannot address.

## Authors’ contributions

RC designed the methods, performed the analysis, and wrote the manuscript. SB conceived the research, designed the methods, and edited the manuscript. AT extracted and processed the data and edited the manuscript.

## Data availability

The primary datasets generated and/or analyzed during this study are not publicly available as they contain proprietary and sensitive health information. However, a sample of the data are available on GitHub.

## Acknowledgments

We gratefully acknowledge Change Healthcare (now part of Optum) for providing claims data and data support. Research reported in this publication was supported by NIGMS of the National Institutes of Health under award number R35GM153478. The content is solely the responsibility of the authors and does not necessarily represent the official views of the sponsors.

## Supplementary Information

### Supplementary Methods

#### National Respiratory and Enteric Virus Surveillance System (NREVSS)

NREVSS monitors virus activity across the United States through a network of laboratories (National Respiratory and Enteric Virus Surveillance System). Participating laboratories voluntarily report the total number of tests conducted for specific viruses as well as the corresponding number of positive tests. The over 450 laboratories include public health, clinical, and commercial testing sites located in all 50 states and the District of Columbia. Viruses covered by the system include influenza, RSV, SARS-CoV-2, among others. These data served as model inputs to account for the time-varying and seasonal nature of influenza. National and state-level test positivity rates were included in the modeling process. To smooth out data noise, a moving average value was calculated from the positive test percent of the current week and 3 preceding weeks.

#### Influenza Hospitalization Surveillance Network (FluSurv-NET)

FluSurv-NET tracks laboratory confirmed influenza hospitalizations among children and adults in the United States (CDC Influenza Hospitalization Surveillance). Data are reported on a weekly basis from a collection of acute care hospitals across 14 states. Hospital catchment areas are estimated to cover over 90 million individuals representing 9 percent of the United States population. Reporting is typically only conducted during the influenza season from October 1 to April 30 every year. To be included in the data, an individual must reside in a participating hospital’s catchment area and test positive for influenza in the 14 days prior to or during hospitalization. This data source served as a comparison to examine trend alignment with model predictions. State-level hospitalization rates were smoothed by calculating a moving average from the hospitalization rate of the current week and 3 preceding weeks.

#### Influenza-Like Illness Surveillance Network (ILI-NET)

ILI-NET surveils outpatient visits for respiratory symptoms that constitute influenza-like illness (CDC Influenza Surveillance). Influenza-like illness is defined as the presence of a fever (temperature of 100 °F or greater) and a cough and/or a sore throat. The broad syndromic definition has potential to measure pathogens that produce a similar set of symptoms such as RSV and SARS-CoV-2. Data is reported weekly throughout the entire year and activity is monitored across all 50 states and the District of Columbia. Over 3,400 health care providers voluntarily report to the system. This data source was used to compare against model predictions. State-level influenza-like illness proportions were smoothed by calculating a moving average from the value of the current week and 3 preceding weeks.

#### New York State (NYS) influenza laboratory-confirmed cases by county

New York State provides weekly counts of laboratory-confirmed influenza at the county level (NYS Influenza Surveillance). Laboratories that test state residents report all positive influenza samples to the health department from October through mid-May of each year, corresponding with the influenza season. Most publicly available disease surveillance sources only included national, regional, or state-level estimates for the period of interest (2016-2020). Therefore, the availability of this data provided a unique opportunity to compare local influenza activity with our syndromic model estimates. County-level case counts were smoothed by calculating a moving average from the cases during the current week and 3 preceding weeks.

#### CDC Influenza Season Burden Estimates

The Centers for Disease Control and Prevention (CDC) provides estimates of influenza burden each season for the entire population and by age group (CDC Influenza Burden). The total number of symptomatic influenza cases across the country, accounting for test sensitivity, testing rates, and healthcare seeking behavior, with corresponding 95% confidence intervals were used to compare to predicted disease burden estimates [33]. The proportion of the country infected with symptomatic influenza each season was calculated by dividing the total number of symptomatic influenza cases by the population for all ages and by age group.

### Wastewater surveillance data

Wastewater data monitoring the concentration of both influenza A and B were included as modeling inputs. Data were provided by WastewaterSCAN, a research-focused wastewater monitoring program operating at over 150 sites across the United States (WastewaterSCAN Surveillance). For our project, we utilized their influenza data from the 2022-23 respiratory virus season. Since wastewater is measured at sewage treatment facilities that cover unique geographic areas, individual sites were assigned to the U.S. county that a majority of their catchment area covered. Daily sampling was aggregated to the weekly scale by taking the mean value for a single site across each week. Finally, to create data at the county-week scale, the mean influenza A and B wastewater concentration across multiple treatment plant locations in a week was calculated for counties with more than one monitoring site.

## Supplementary Figures & Tables

**Table S1:** ICD-10 codes used to define influenza and respiratory syncytial virus (RSV) confirmed infection.

| Disease | Code | Description |
| --- | --- | --- |
| Influenza | J09.X1 | Influenza due to identified novel influenza A virus with pneumonia |
| Influenza | J09.X2 | Influenza due to identified novel influenza A virus with other respiratory manifestations |
| Influenza | J09.X3 | Influenza due to identified novel influenza A virus with gastrointestinal manifestations |
| Influenza | J09.X9 | Influenza due to identified novel influenza A virus with other manifestations |
| Influenza | J10.00 | Influenza due to other identified influenza virus with unspecified type of pneumonia |
| Influenza | J10.01 | Influenza due to other identified influenza virus with the same other identified influenza virus pneumonia |
| Influenza | J10.08 | Influenza due to other identified influenza virus with other specified pneumonia |
| Influenza | J10.1 | Influenza due to other identified influenza virus with other respiratory manifestations |
| Influenza | J10.2 | Influenza due to other identified influenza virus with gastrointestinal manifestations |
| Influenza | J11.81 | Influenza due to other identified influenza virus with encephalopathy |
| Influenza | J11.82 | Influenza due to other identified influenza virus with myocarditis |
| Influenza | J11.83 | Influenza due to other identified influenza virus with otitis media |
| Influenza | J11.89 | Influenza due to other identified influenza virus with other manifestations |
| Influenza | J11.00 | Influenza due to unidentified influenza virus with unspecified type of pneumonia |
| Influenza | J11.08 | Influenza due to unidentified influenza virus with specified pneumonia |
| Influenza | J11.1 | Influenza due to unidentified influenza virus with other respiratory manifestations |
| Influenza | J11.2 | Influenza due to unidentified influenza virus with gastrointestinal manifestations |
| Influenza | J11.81 | Influenza due to unidentified influenza virus with encephalopathy |
| Influenza | J11.82 | Influenza due to unidentified influenza virus with myocarditis |
| Influenza | J11.83 | Influenza due to unidentified influenza virus with otitis media |
| Influenza | J11.89 | Influenza due to unidentified influenza virus with other manifestations |
| RSV | B97.4 | Respiratory syncytial virus as the cause of diseases classified elsewhere |
| RSV | J12.1 | Respiratory syncytial virus pneumonia |
| RSV | J20.5 | Acute bronchitis due to respiratory syncytial virus |
| RSV | J21.0 | Acute bronchiolitis due to respiratory syncytial virus |

**Table S2:**
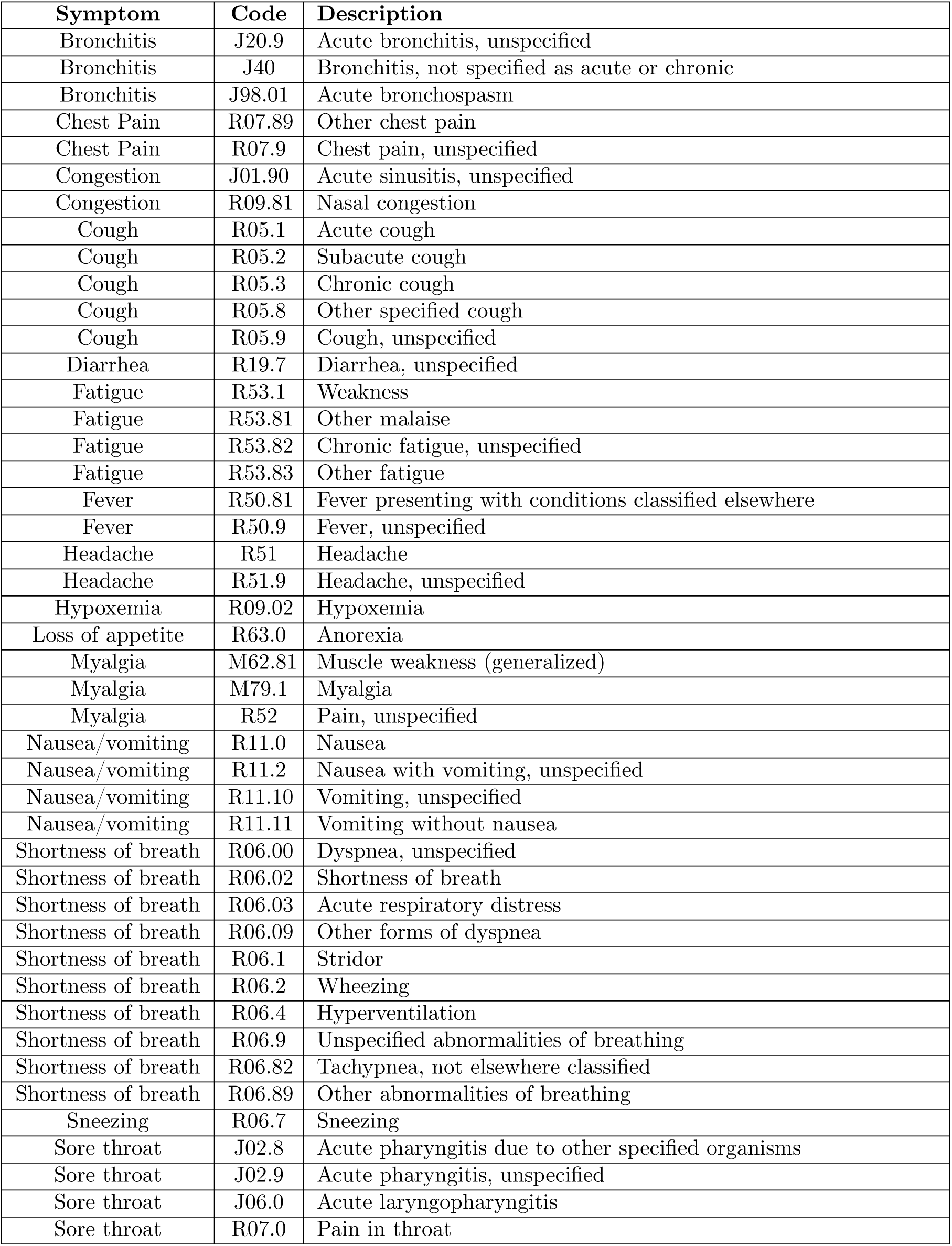
ICD-10 codes used to define the 15 signs and symptoms used in the analysis.

| Symptom | Code | Description |
| --- | --- | --- |
| Bronchitis | J20.9 | Acute bronchitis, unspecified |
| Bronchitis | J40 | Bronchitis, not specified as acute or chronic |
| Bronchitis | J98.01 | Acute bronchospasm |
| Chest Pain | R07.89 | Other chest pain |
| Chest Pain | R07.9 | Chest pain, unspecified |
| Congestion | J01.90 | Acute sinusitis, unspecified |
| Congestion | R09.81 | Nasal congestion |
| Cough | R05.1 | Acute cough |
| Cough | R05.2 | Subacute cough |
| Cough | R05.3 | Chronic cough |
| Cough | R05.8 | Other specified cough |
| Cough | R05.9 | Cough, unspecified |
| Diarrhea | R19.7 | Diarrhea, unspecified |
| Fatigue | R53.1 | Weakness |
| Fatigue | R53.81 | Other malaise |
| Fatigue | R53.82 | Chronic fatigue, unspecified |
| Fatigue | R53.83 | Other fatigue |
| Fever | R50.81 | Fever presenting with conditions classified elsewhere |
| Fever | R50.9 | Fever, unspecified |
| Headache | R51 | Headache |
| Headache | R51.9 | Headache, unspecified |
| Hypoxemia | R09.02 | Hypoxemia |
| Loss of appetite | R63.0 | Anorexia |
| Myalgia | M62.81 | Muscle weakness (generalized) |
| Myalgia | M79.1 | Myalgia |
| Myalgia | R52 | Pain, unspecified |
| Nausea/vomiting | R11.0 | Nausea |
| Nausea/vomiting | R11.2 | Nausea with vomiting, unspecified |
| Nausea/vomiting | R11.10 | Vomiting, unspecified |
| Nausea/vomiting | R11.11 | Vomiting without nausea |
| Shortness of breath | R06.00 | Dyspnea, unspecified |
| Shortness of breath | R06.02 | Shortness of breath |
| Shortness of breath | R06.03 | Acute respiratory distress |
| Shortness of breath | R06.09 | Other forms of dyspnea |
| Shortness of breath | R06.1 | Stridor |
| Shortness of breath | R06.2 | Wheezing |
| Shortness of breath | R06.4 | Hyperventilation |
| Shortness of breath | R06.9 | Unspecified abnormalities of breathing |
| Shortness of breath | R06.82 | Tachypnea, not elsewhere classified |
| Shortness of breath | R06.89 | Other abnormalities of breathing |
| Sneezing | R06.7 | Sneezing |
| Sore throat | J02.8 | Acute pharyngitis due to other specified organisms |
| Sore throat | J02.9 | Acute pharyngitis, unspecified |
| Sore throat | J06.0 | Acute laryngopharyngitis |
| Sore throat | R07.0 | Pain in throat |

**Figure S1:**
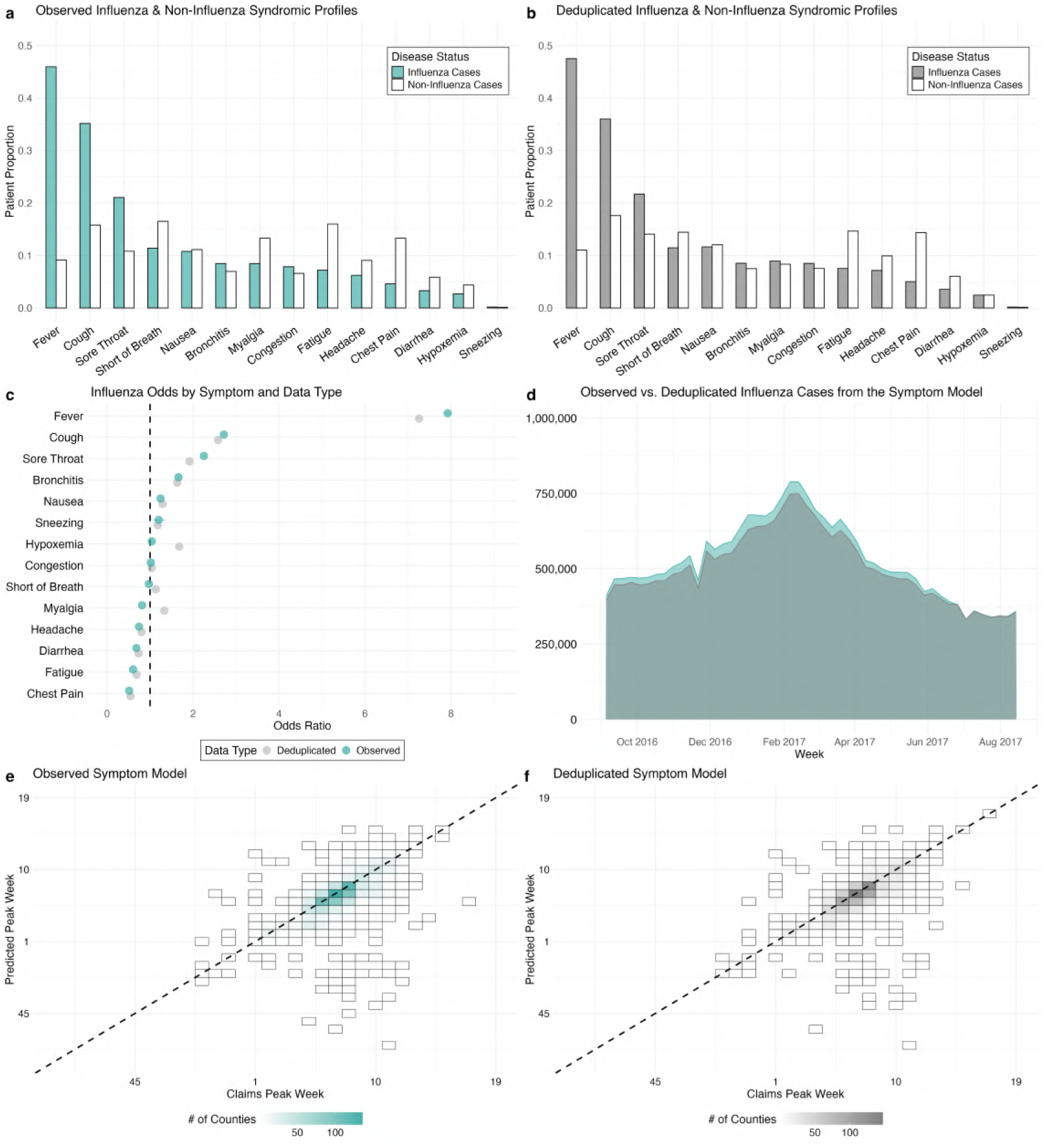
Descriptive and modeling differences between observed and deduplicated data. (**a**) Proportion of influenza cases and non-influenza cases with 14 different symptoms from observed data. (**b**) Proportion of influenza cases and non-influenza cases with 14 different symptoms from deduplicated data. (**c**) Logistic regression results displaying the odds ratios of influenza given symptom presence, for observed and deduplicated data. A vast majority of odds ratios were qualitatively similar between the two data types. Both proportions and regression results were produced from balanced samples of cases and non-cases from 2016-2020, representing a subset of the full medical claims data source. Odds ratios produced by each data type were from the Symptom model, where confirmed disease was the dependent variable and all symptoms served as independent predictors.(**d**) National predicted syndromic influenza cases produced by the Symptom model from August 2016 to July 2017 for both observed and deduplicated data. (**e**) Syndromic predicted peak time from the Symptom model with observed data compared to confirmed influenza peak time from the claims data.(**f**) Syndromic predicted peak time from the Symptom model with deduplicated data compared to confirmed influenza peak time from the claims data. Spearman correlation coefficients and root mean square error (RMSE) values were 0.67 vs. 0.68 and 17.8 vs. 16.1 days for the observed vs. deduplicated data models, respectively.

**Figure S2:**
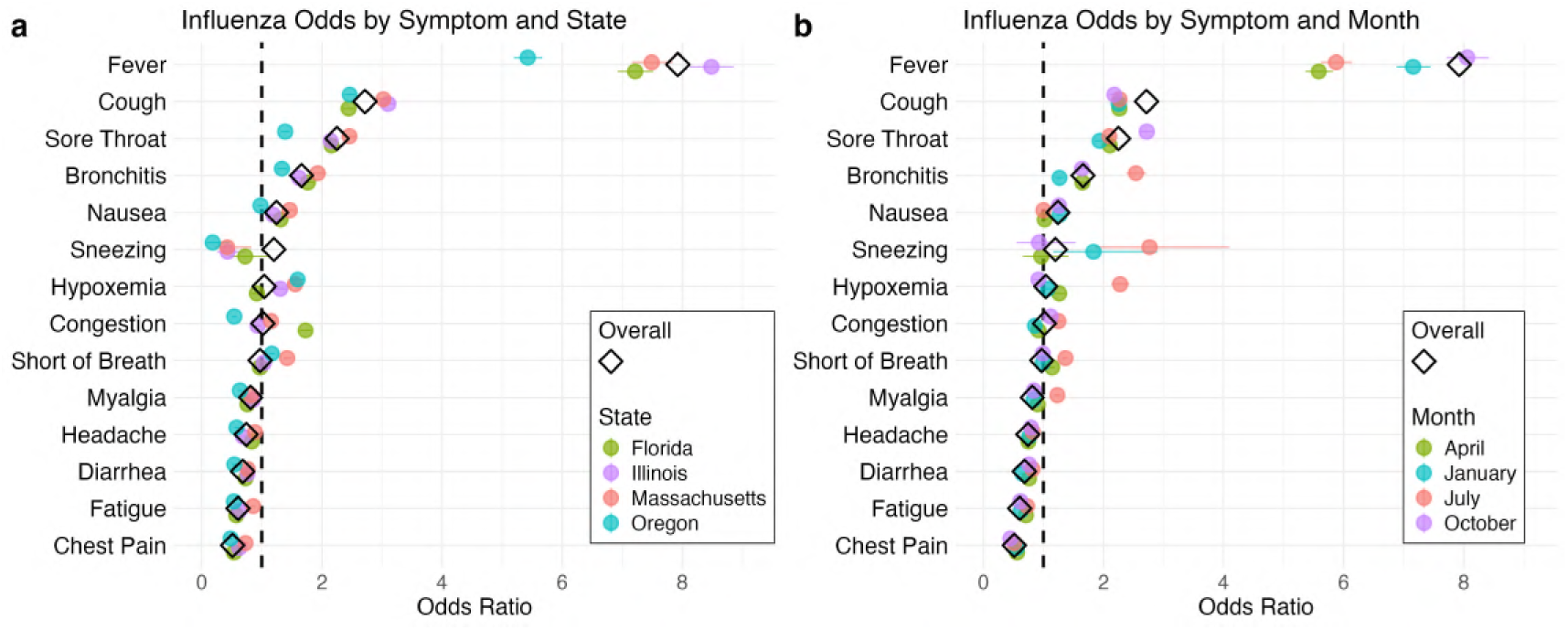
Influenza odds by symptom, geography, and time. (**a**) Logistic regression results displaying the odds ratios of influenza given symptom presence, across the entire country and for four states: Florida, Illinois, Massachusetts, and Oregon. (**b**) Logistic regression results displaying the odds ratios of influenza given symptom presence, across all months and for four specific months: January, April, July, and October. Regression results were produced from balanced samples of cases and non-cases from 2016-2020, representing a subset of the full medical claims data source. Odds ratios produced by each data type were from the Symptom model, where confirmed disease was the dependent variable and all symptoms served as independent predictors.

**Figure S3:**
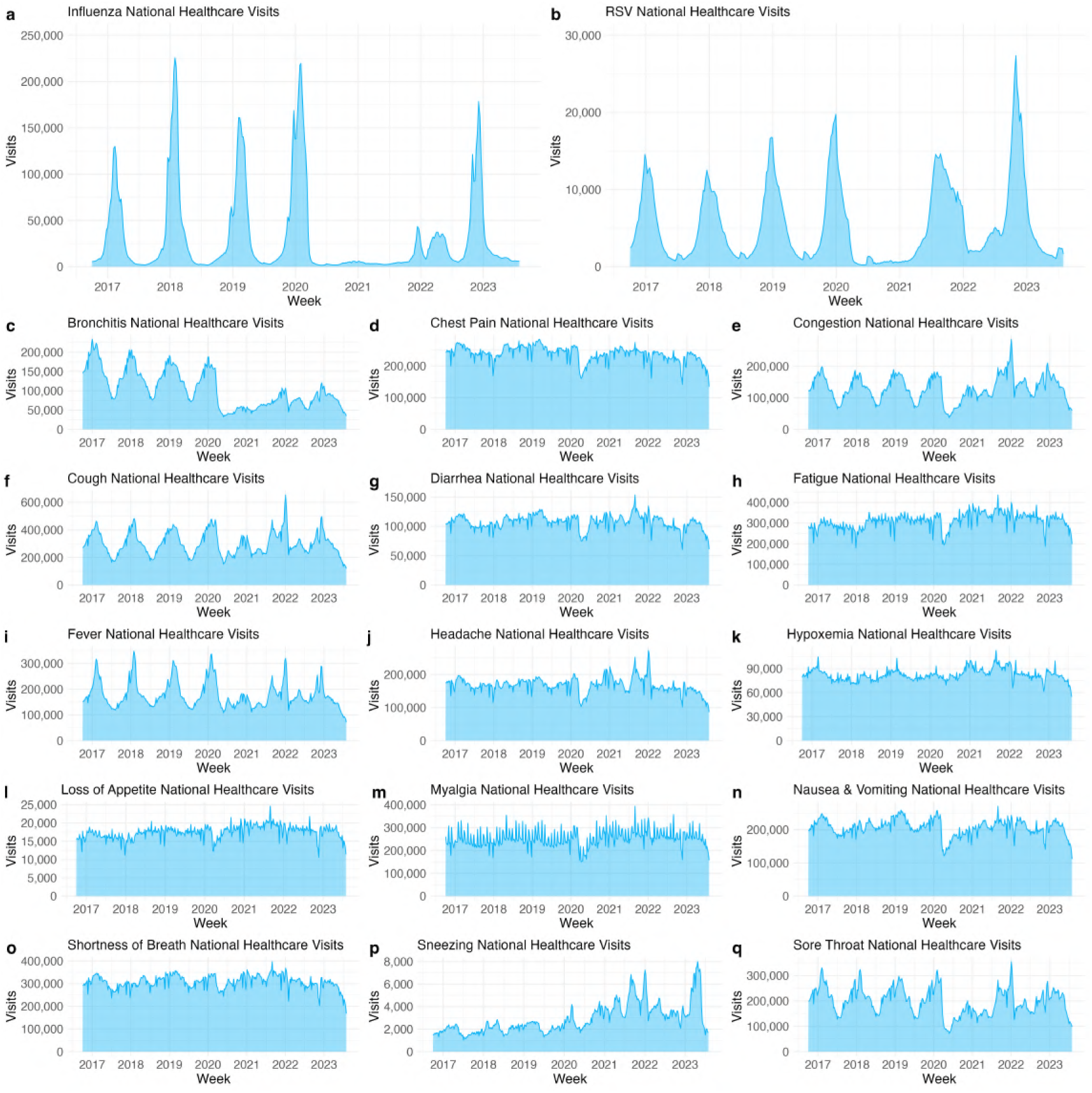
Disease and symptom national observed temporal trends, 2016-2023. (**a**) Confirmed influenza healthcare visits in the United States. (**b**) Confirmed RSV healthcare visits in the United States. (**c-q**) United States healthcare visits where bronchitis, chest pain, congestion, cough, diarrhea, fatigue, fever, headache, hypoxemia, loss of appetite, myalgia, nausea/vomiting, shortness of breath, sneezing, or sore throat was indicated as a symptom.

**Figure S4:**
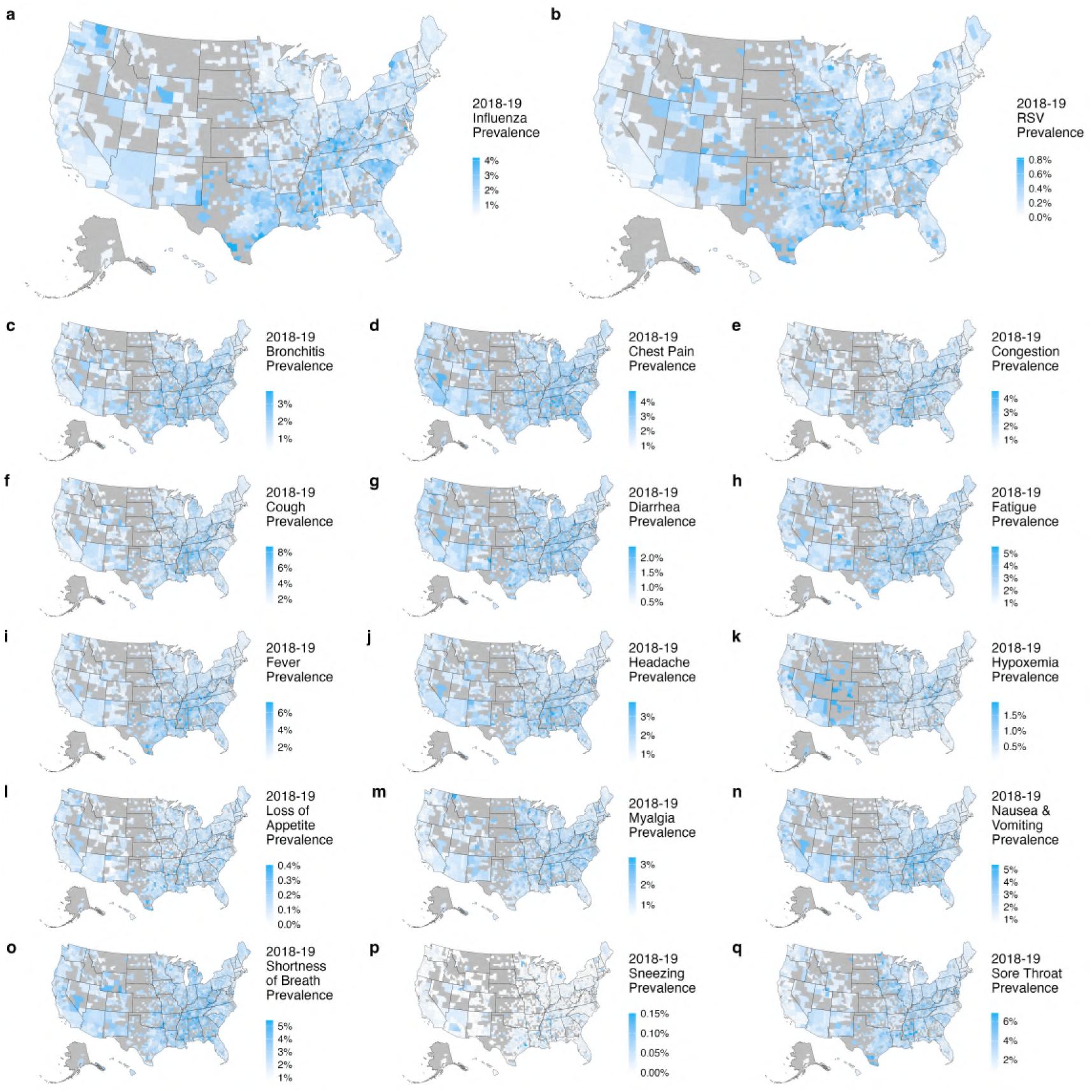
Disease and symptom county-level observed prevalence, 2018-2019. (**a**) County-level confirmed influenza prevalence for the 2018-19 respiratory virus season. (**b**) County-level confirmed RSV prevalence for the 2018-19 respiratory virus season. (**c-q**) County-level bronchitis, chest pain, congestion, cough, diarrhea, fatigue, fever, headache, hypoxemia, loss of appetite, myalgia, nausea/vomiting, shortness of breath, sneezing, or sore throat prevalence for the 2018-19 respiratory virus season. Prevalence estimates were calculated by dividing the deduplicated number of disease or symptom cases by the total number of unique patients (all causes) in each county from October 2018 through April 2019. Counties with small population sizes (< 10,000), low numbers of claims (< 5,000), or with outlier values (> 3 standard deviations above the mean) were excluded from the prevalence estimates.

**Figure S5:**
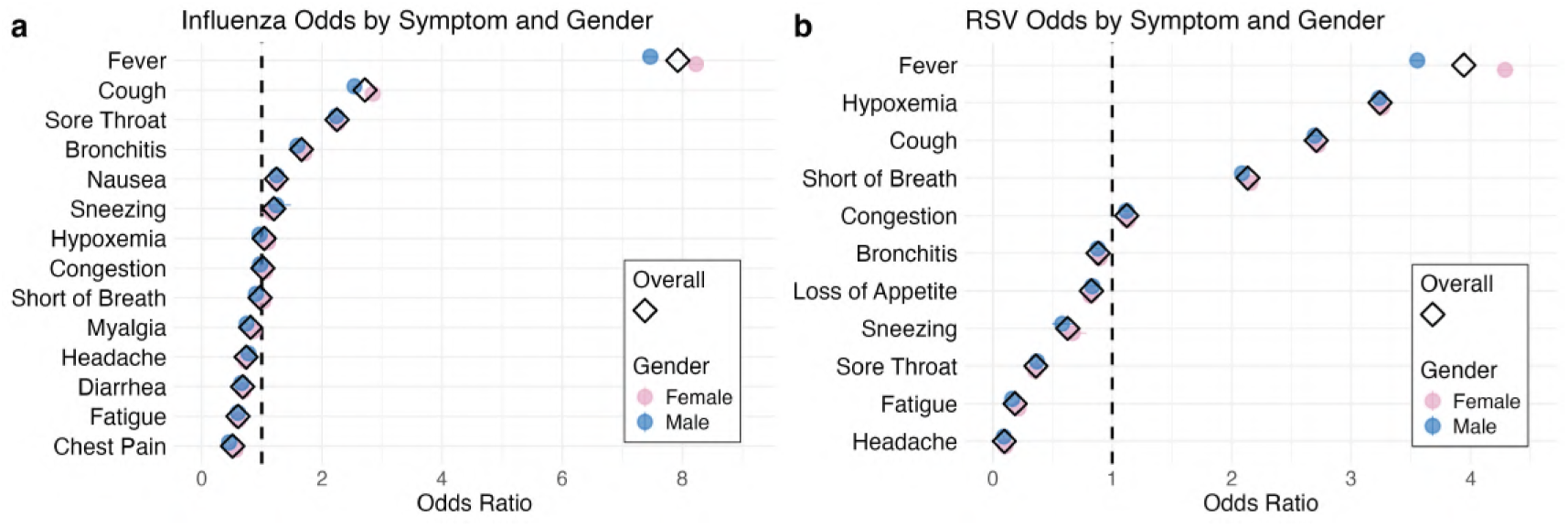
Disease odds by symptom and gender for influenza and RSV. (**a**) Logistic regression results displaying the odds ratios of influenza given symptom presence, for the total population and by gender. (**b**) Logistic regression results displaying the odds ratios of RSV given symptom presence, for the total population and by gender. Regression results were produced from balanced samples of cases and non-cases from 2016-2020, representing a subset of the full medical claims data source. Overall and gender-specific odds ratios were from the Symptom Model, where confirmed disease was the dependent variable and all symptoms served as independent predictors.

**Figure S6:**
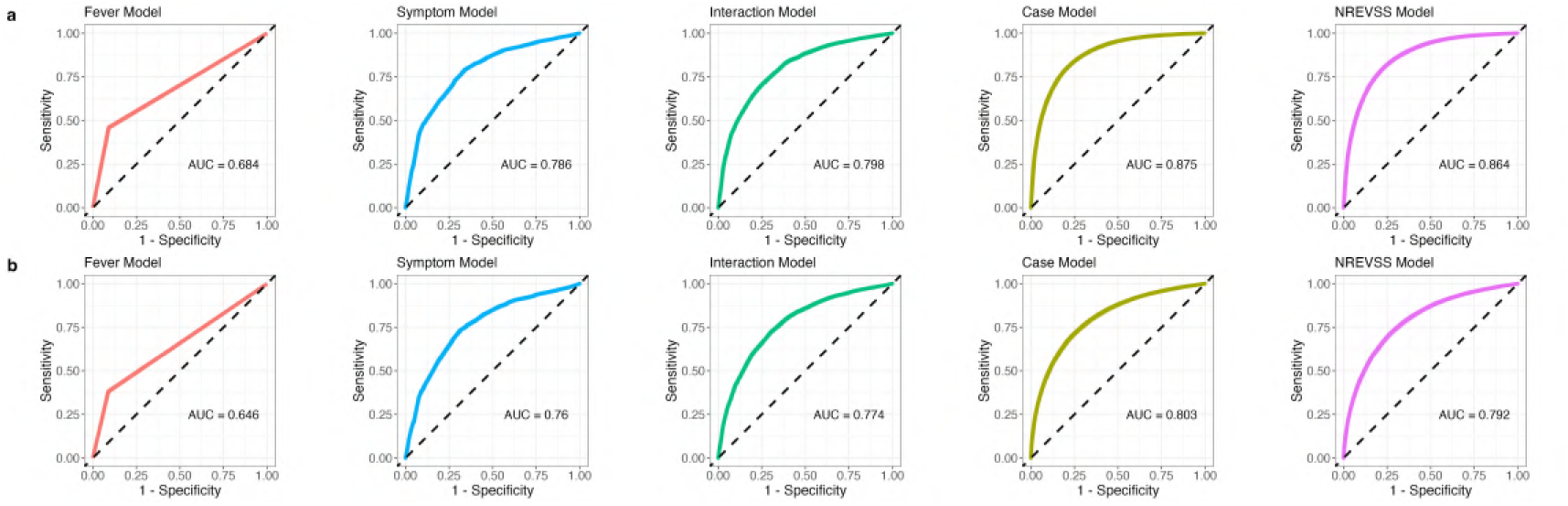
Receiver Operating Characteristic (ROC) curves and AUC values by influenza model. (**a**) ROC curves and AUC values predicting influenza disease status with medical claims from 2016-2020, by model. (**b**) ROC curves and AUC values predicting influenza disease status with medical claims from 2021-2023, by model. The Wastewater Model was not included in this analysis since only data from 2022-2023 was used to train the model.

**Figure S7:**
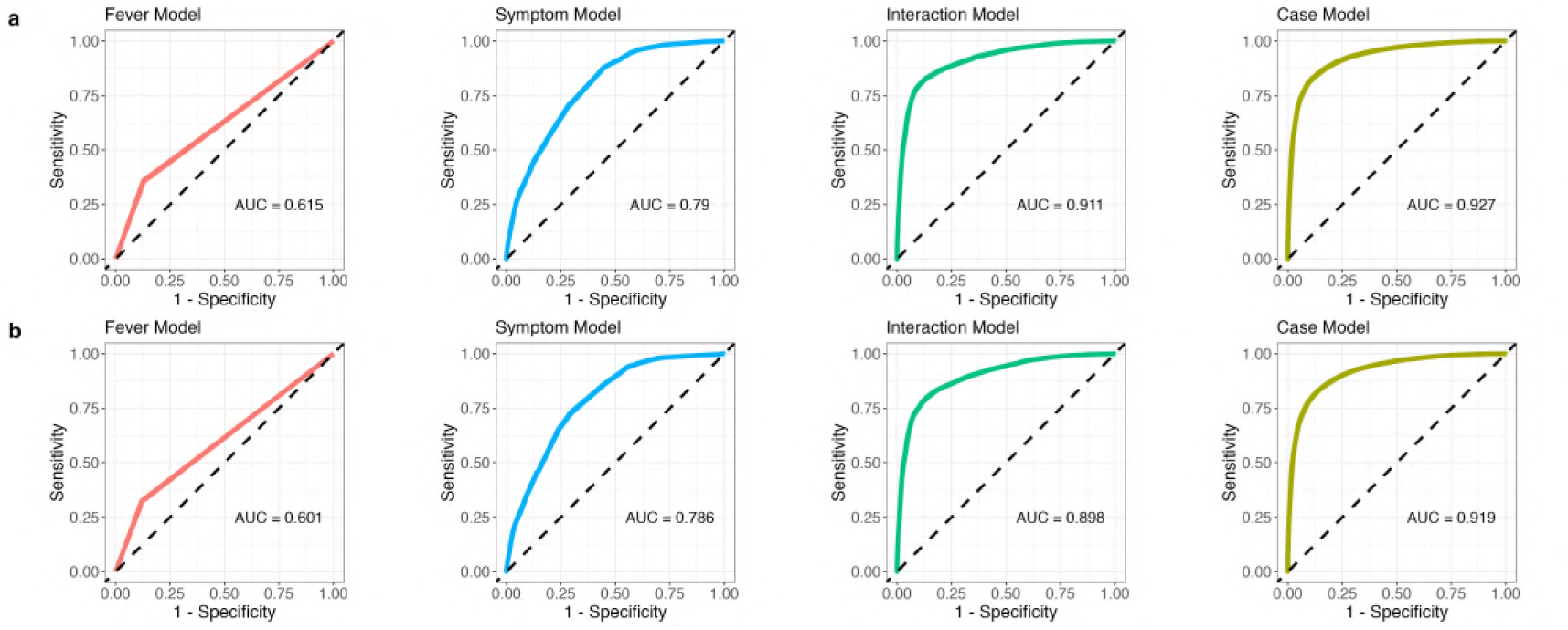
Receiver Operating Characteristic (ROC) curves and AUC values by RSV model. (**a**) ROC curves and AUC values predicting RSV disease status with medical claims from 2016-2020, by model. (**b**) ROC curves and AUC values predicting RSV disease status with medical claims from 2021-2023, by model.

**Figure S8:**
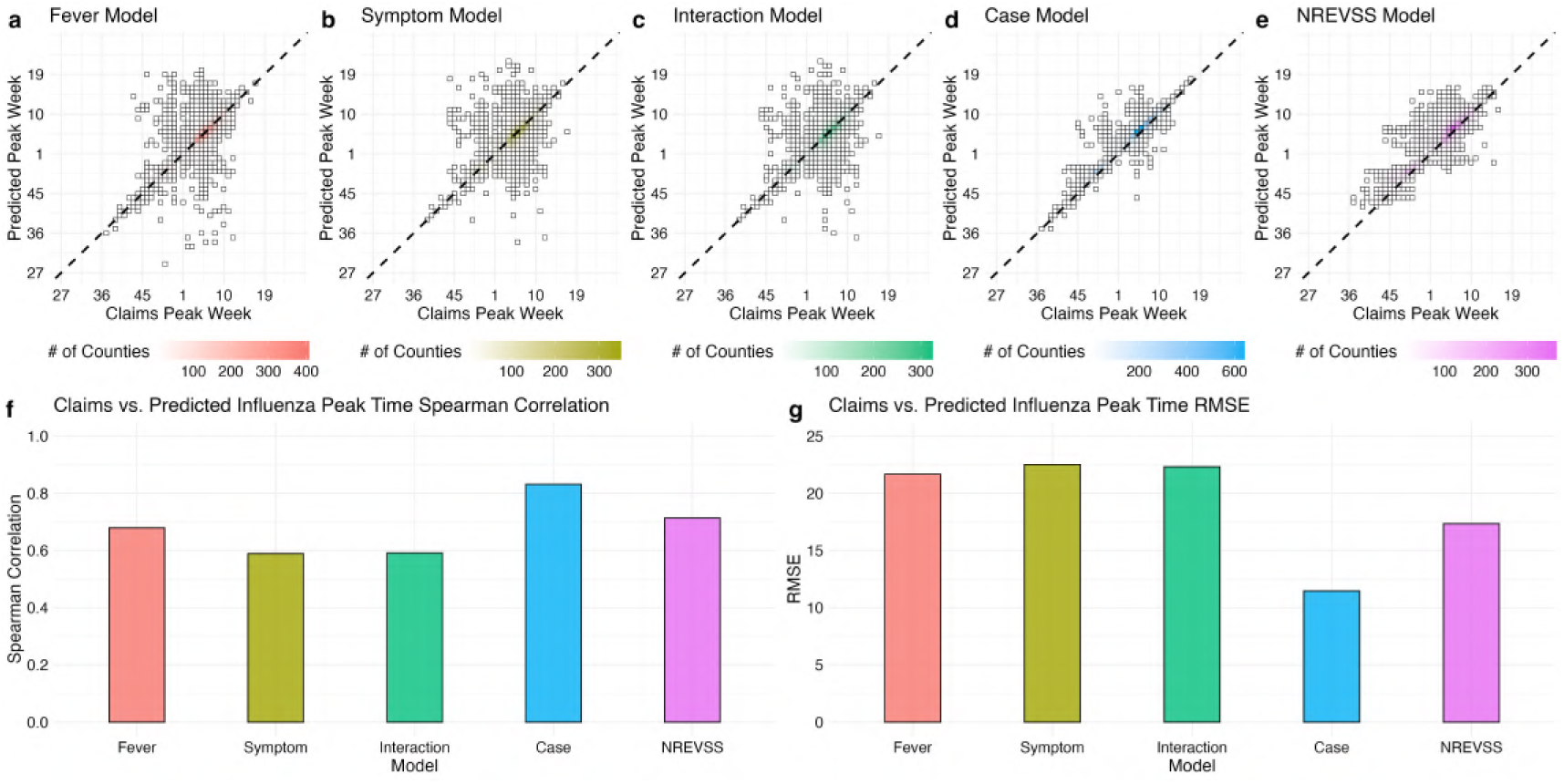
Influenza peak time validation by model. (**a**) Syndromic predicted peak time from the Fever model compared to confirmed influenza peak time from the claims data. (**b**) Syndromic predicted peak time from the Symptom model compared to confirmed influenza peak time from the claims data. (**c**) Syndromic predicted peak time from the Interaction model compared to confirmed influenza peak time from the claims data. (**d**) Syndromic predicted peak time from the Case model compared to confirmed influenza peak time from the claims data. (**e**) Syndromic predicted peak time from the NREVSS model compared to confirmed influenza peak time from the claims data. (**f**) Spearman correlation coefficients between predicted peak and claims peak by model across five respiratory virus seasons (2016-17, 2017-18, 2018-19, 2019-20, 2022-23). (**g**) Root mean square error (RMSE) between predicted peak and claims peak by model across the same five respiratory virus seasons. The 2020-21 season was excluded due to low influenza circulation during the COVID-19 pandemic while the 2021-22 season was excluded due to the occurrence of unusual multi-peak influenza dynamics.

**Figure S9:**
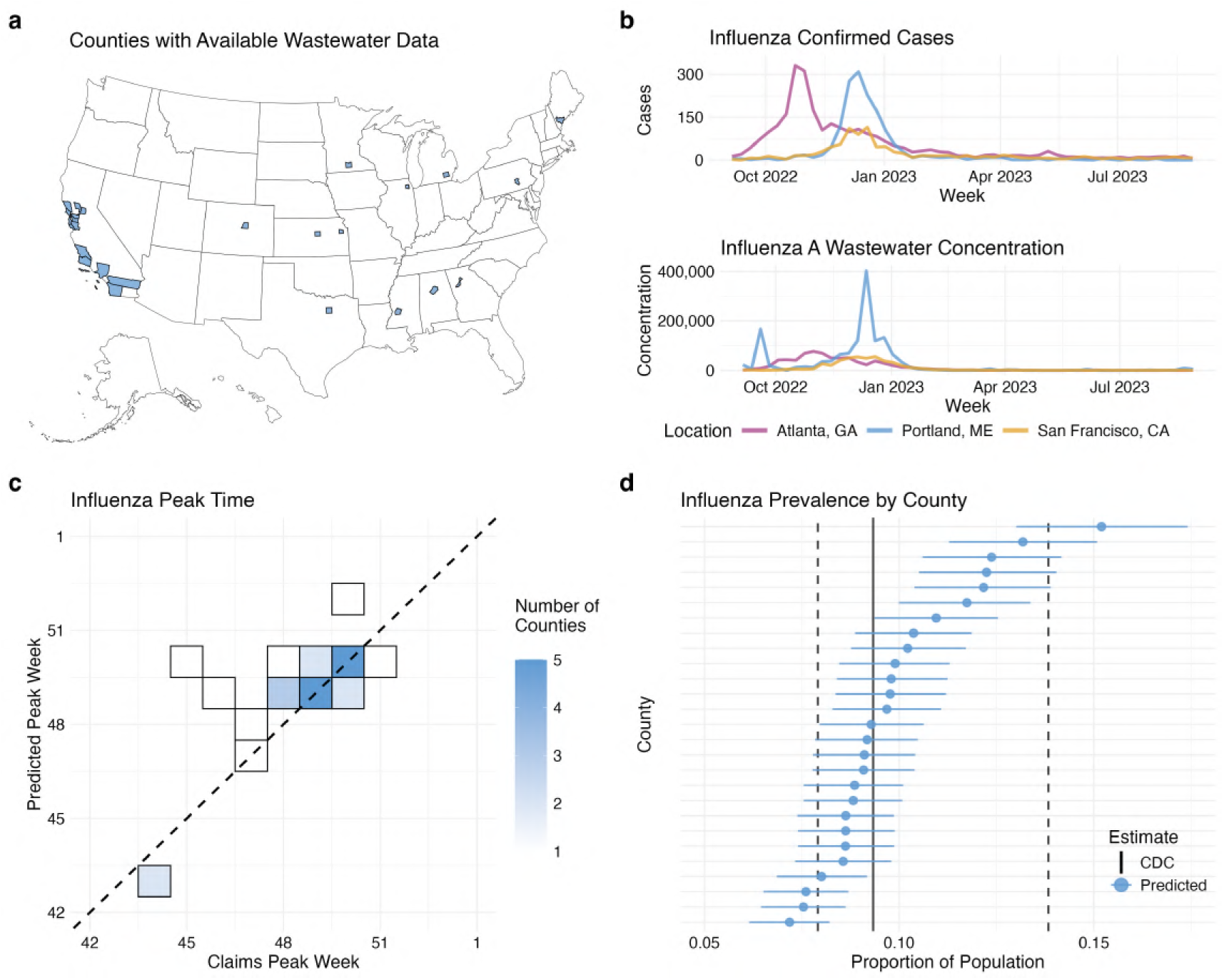
Using Wastewater Data to Compliment Syndromic Surveillance. (**a**) The 27 counties with available and complete wastewater samples during the 2022-23 respiratory virus season. (**b**) Confirmed influenza cases from medical claims data compared against influenza A concentrations measured in wastewater from three select locations: Atlanta, GA, Portland, ME, and San Francisco, CA. (**c**) County peak time correlation between predicted influenza and confirmed influenza during the 2022-23 season. (**d**) County-level influenza prevalence estimates with corresponding 95% confidence intervals. The black solid line displays the CDC national influenza prevalence estimate while dashed lines are the 95% confidence range for the estimate.

**Figure S10:**
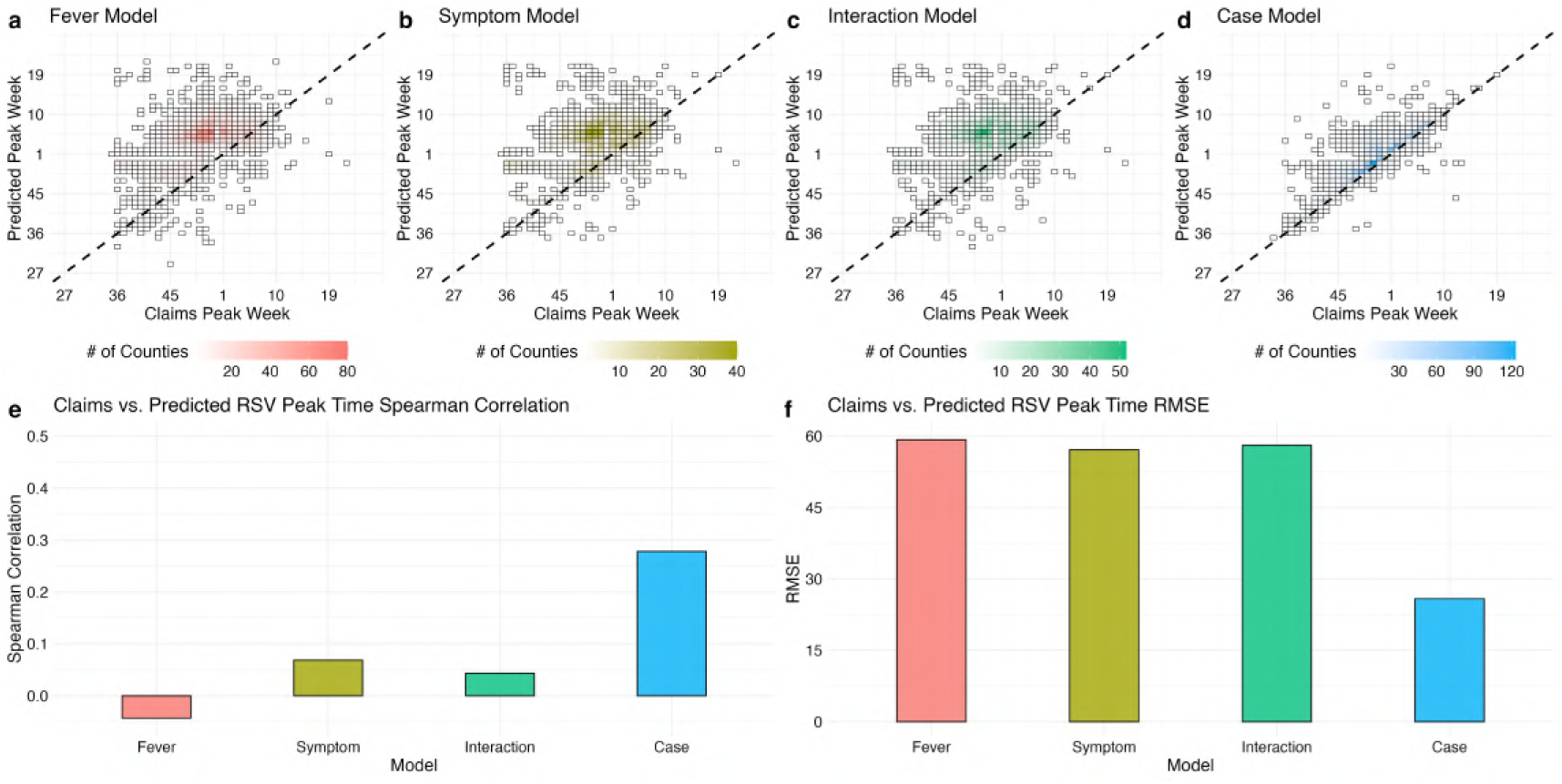
RSV peak time validation by model. (**a**) Syndromic predicted peak time from the Fever model compared to confirmed RSV peak time from the claims data. (**b**) Syndromic predicted peak time from the Symptom model compared to confirmed RSV peak time from the claims data. (**c**) Syndromic predicted peak time from the Interaction model compared to confirmed RSV peak time from the claims data. (**d**) Syndromic predicted peak time from the Case model compared to confirmed RSV peak time from the claims data. (**f**) Spearman correlation coefficients between predicted peak and claims peak by model across five respiratory virus seasons (2016-17, 2017-18, 2018-19, 2019-20, 2021-22, 2022-23). (**g**) Root mean square error (RMSE) between predicted peak and claims peak by model across the same five respiratory virus seasons. The 2020-21 and 2021-22 seasons were excluded due to low RSV circulation during the COVID-19 pandemic.

**Figure S11:**
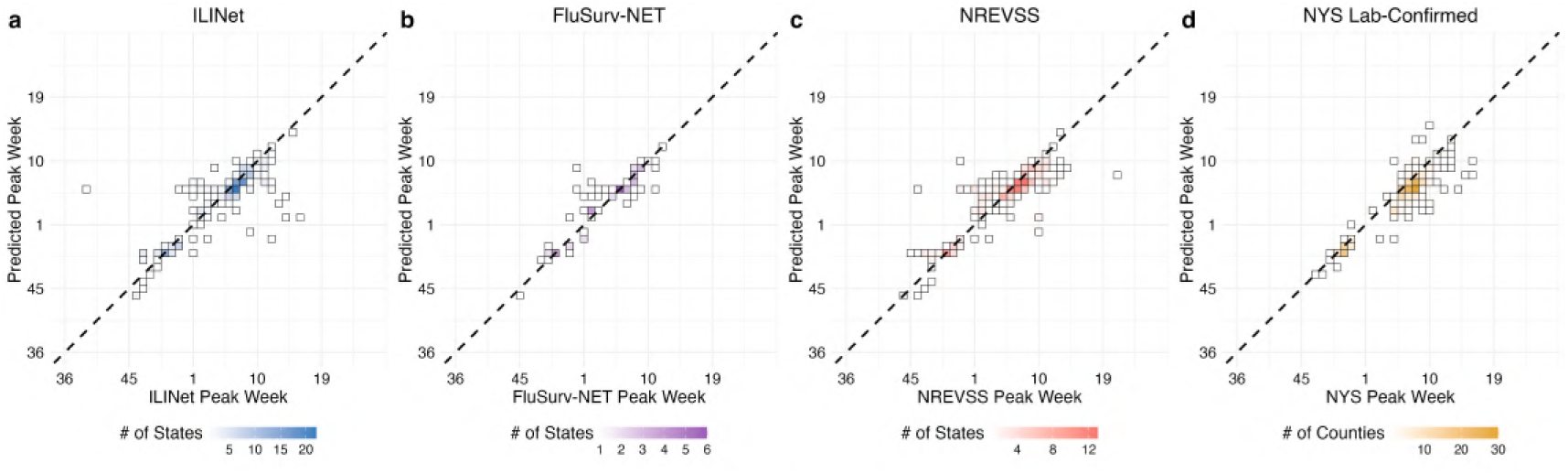
External Influenza peak time validation for the Case Model. (**a**) Syndromic predicted peak time from the Case model compared to ILINet surveillance data at the state-level. (**b**) Syndromic predicted peak time from the Case model compared to FluSurv-NET surveillance data at the state-level. (**c**) Syndromic predicted peak time from the Case model compared to NREVSS surveillance data at the state-level. (**d**) Syndromic predicted peak time from the Case model compared to New York State lab-confirmed surveillance data at the county-level. Scatter plots show data across five respiratory virus seasons (2016-17, 2017-18, 2018-19, 2019-20, 2022-23). The 2020-21 season was excluded due to low influenza circulation during the COVID-19 pandemic while the 2021-22 season was excluded due to the occurrence of unusual multi-peak dynamics.

**Figure S12:**
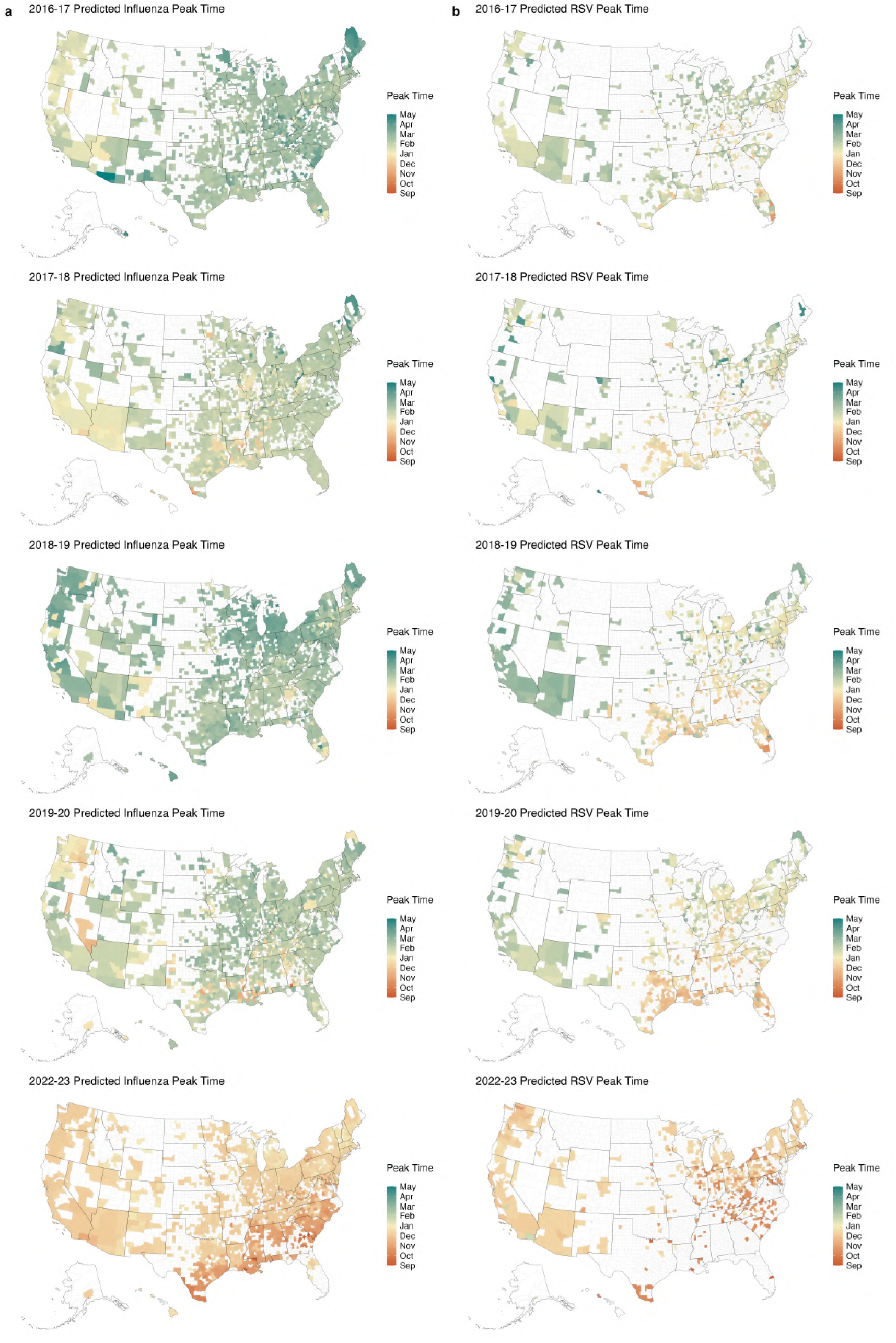
County-level Syndromic peak time predictions for Influenza and RSV across Seasons. (**a**) Syndromic influenza peak time predictions from the Case Model across five respiratory virus seasons (2016-17, 2017-18, 2018-19, 2019-20, 2022-23). (**b**) Syndromic RSV peak time predictions from the Case Model across five respiratory virus seasons (2016-17, 2017-18, 2018-19, 2019-20, 2022-23). White counties either represent areas with fewer than 10,000 inhabitants, fever than 5,000 total claims during the season, or where no peak was identified.

**Figure S13:**
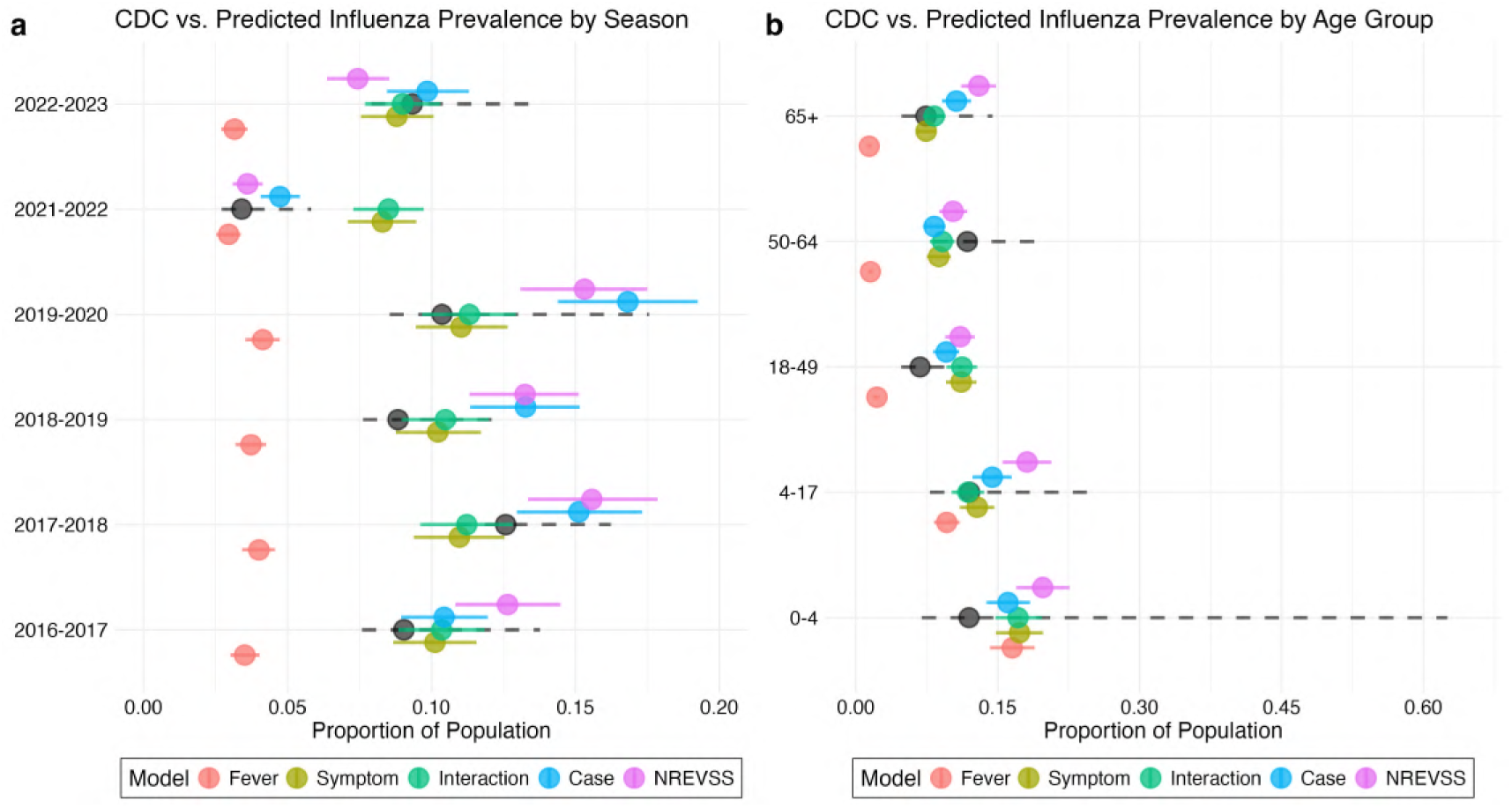
Predicted Syndromic Influenza Prevalence vs. CDC across Seasons and Age Groups. (**a**) **a**) National influenza prevalence for the entire population comparing the CDC estimate to the syndromic prediction across seasons, by model. (**b**) National influenza prevalence for the 2016-17 season comparing the CDC estimate to the syndromic prediction across age groups, by model. Black circles and dashed lines represent the CDC estimate with corresponding 95% confidence interval. The 2020-21 influenza season was excluded from this analysis due to low virus circulation.

**Figure S14:**
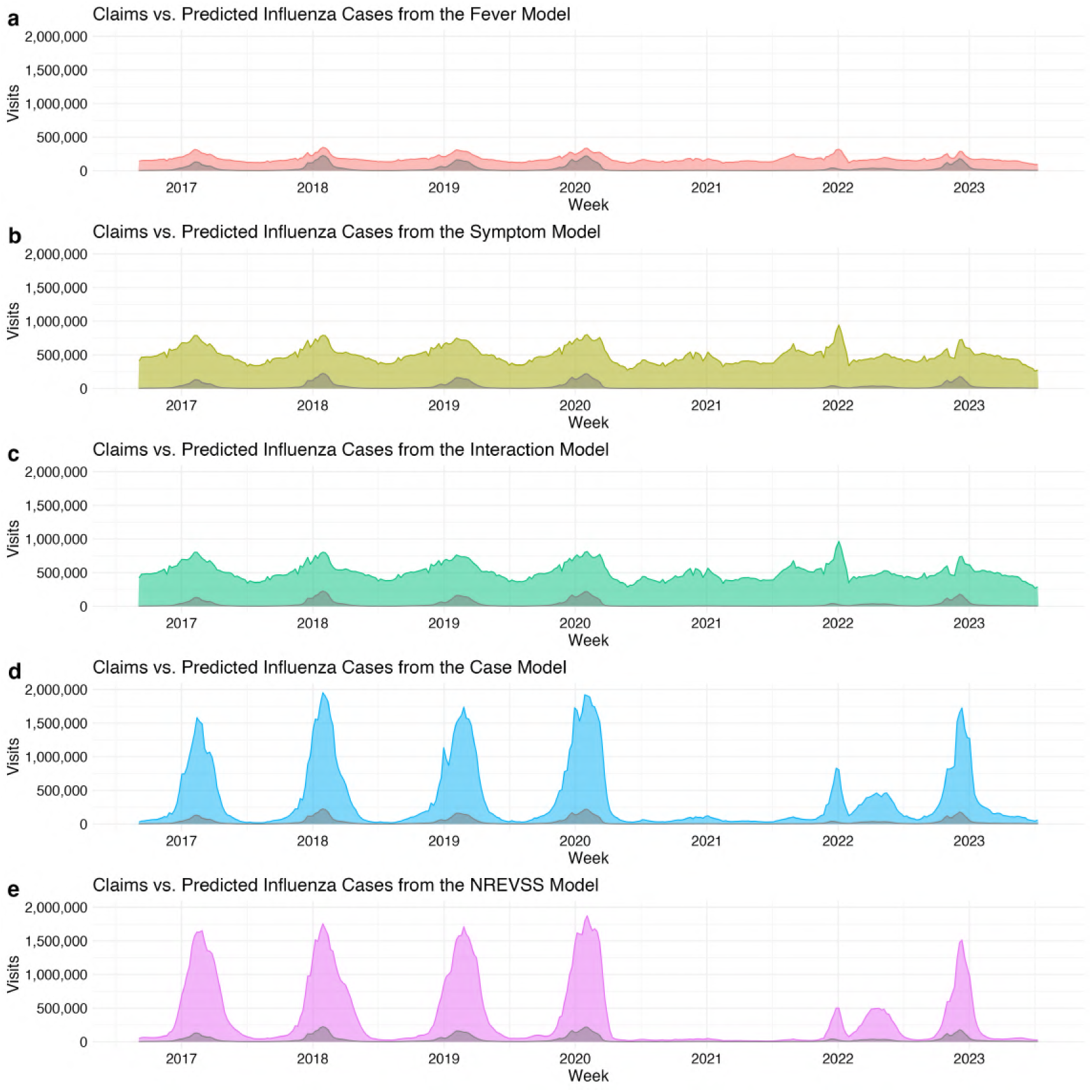
National Predicted Syndromic Influenza Cases over Time by Model. **(a)** National predicted syndromic influenza cases from the Fever Model from August 2016 to July 2023. (**b**) National predicted syndromic influenza cases from the Symptom Model from August 2016 to July 2023. (**c**) National predicted syndromic influenza cases from the Interaction Model from August 2016 to July 2023. (**d**) National predicted syndromic influenza cases from the Case Model from August 2016 to July 2023. (**e**) National predicted syndromic influenza cases from the NREVSS Model from August 2016 to July 2023. The dark shaded area in each plot represents confirmed influenza cases from the medical claims data. Predicted cases display results from models trained on data during the 2016-2020 time frame, so all values after August 2020 are out-of-sample predictions.

